# Potential Effectiveness and Safety of Antiviral Agents in Children with Coronavirus Disease 2019: A Rapid Review and Meta-Analysis

**DOI:** 10.1101/2020.04.13.20064436

**Authors:** Qianling Shi, Qi Zhou, Xia Wang, Jing Liao, Yang Yu, Zijun Wang, Shuya Lu, Yanfang Ma, Yangqin Xun, Xufei Luo, Weiguo Li, Toshio Fukuoka, Hyeong Sik Ahn, Myeong Soo Lee, Zhengxiu Luo, Enmei Liu, Yaolong Chen, Qubei Li, Kehu Yang, on behalf of COVID-19 evidence and recommendations working group

**Affiliations:** The First School of Clinical Medicine, Lanzhou University, Lanzhou 730000, China; Evidence-based Medicine Center, School of Basic Medical Sciences, Lanzhou University, Lanzhou 730000, China; Department of Respiratory Medicine, Children’s Hospital of Chongqing Medical University, Chongqing 400014, China; National Clinical Research Center for Child Health and Diseases, Ministry of Education Key Laboratory of Child Development and Disorders, China International Science and Technology Cooperation Base of Child Development and Critical Disorders, Children’s Hospital of Chongqing Medical University, Chongqing 400014, China; Chongqing Key Laboratory of Pediatrics, Chongqing 400014, China; The Second School of Clinical Medicine, Lanzhou University, Lanzhou 730000, China; Department of Pediatric, Sichuan Provincial People’s Hospital, University of Electronic Science and Technology of China, Chengdu 611731, China; Chinese Academy of Sciences Sichuan Translational Medicine Research Hospital, Chengdu 610072, China; School of Public Health, Lanzhou University, Lanzhou 730000, China; Emergency and Critical Care Center, the Department of General Medicine, Department of Research and Medical Education at Kurashiki Central Hospital, Japan; Advisory Committee in Cochrane Japan, Japan; Department of Preventive Medicine, Korea University College of Medicine, Seoul, Korea; Korea Cochrane Centre, Korea; Korea Institute of Oriental Medicine, Daejeon, Korea; University of Science and Technology, Daejeon, Korea; Lanzhou University, an Affiliate of the Cochrane China Network, Lanzhou 730000, China; Chinese GRADE Center, Lanzhou 730000, China; Key Laboratory of Evidence Based Medicine and Knowledge Translation of Gansu Province, Lanzhou University, Lanzhou 730000, China

**Author notes:** **Correspondence to:** Kehu Yang. Department of the First School of Clinical Medicine, Lanzhou University, Lanzhou 730000, China.; Qubei Li. Department of the Respiratory Medicine, Children’s Hospital of Chongqing Medical University, Chongqing 400014, China. These authors contributed equally to this work (Qianling Shi, Qi Zhou, Xia Wang).

**Keywords:** Antiviral agents, children, COVID-19, meta-analysis, rapid review

## Abstract

**Background:** The COVID-19 outbreak presents a new, life-threatening disease. Our aim was to assess the potential effectiveness and safety of antiviral agents for COVID-19 in children.

**Methods:** Electronic databases from their inception to March, 31 2020 were searched for randomized controlled trials, clinical controlled trials and cohort studies of interventions with antiviral agents for children (less than 18 years of age) with COVID-19.

**Results:** A total of 23 studies of indirect evidence with 6008 patients were included. The risks of bias in all studies were moderate to high in general. The effectiveness and safety of antiviral agents for children with COVID-19 is uncertain: For adults with COVID-19, lopinavir/ritonavir had no effect on mortality (risk ratio [RR]= 0.77, 95% confidence interval [CI] 0.45 to 1.30) and probability of negative PCR test (RR=0.98, 95 CI% 0.82 to 1.18). Arbidol had no benefit on probability of negative PCR test (RR=1.27, 95% CI 0.93 to 1.73). Hydroxychloroquine was not associated with increasing the probability of negative PCR result (RR=0.93, 95% CI 0.73 to 1.18). For adults with SARS, interferon was associated with reduced corticosteroid dose (weighted mean difference [WMD]=-0.14 g, 95% CI -0.21 to -0.07) but had no effect on mortality (RR=0.72, 95% CI 0.28 to 1.88); ribavirin did not reduce mortality (RR=0.68, 95% CI % 0.43 to 1.06) and was associated with high risk of severe adverse reactions; and oseltamivir had no effect on mortality (RR=0.87, 95% CI 0.55 to 1.38). Ribavirin combined with interferon was also not effective in adults with MERS and associated with adverse reactions.

**Conclusions:** There is no evidence showing the effectiveness of antiviral agents for children with COVID-19, and the clinical efficacy of existing antiviral agents is still uncertain. We do not suggest clinical routine use of antivirals for COVID-19 in children, with the exception of clinical trials.

## Background

A novel coronavirus, later named as severe acute respiratory syndrome coronavirus 2 (SARS-CoV-2), was first detected on December 8, 2019, when several cases of pneumonia of unknown etiology were reported in Wuhan, Hubei province, China. (1-3) Due to the rapidly increasing numbers of infections and deaths, World Health Organization (WHO) subsequently declared the outbreak as a Public Health Emergency of International Concern (PHEIC) on January 30, 2019 and officially named the disease as “Corona Virus Disease hyphen one nine” (COVID-19) on February 11, 2020.(4-6) As of April 12, a total of 1,696,588 confirmed cases had been reported in more than 200 countries, and the number of cases abroad was still rapidly increasing, creating global alarm and concerns about the impact on health care and economy of the affected areas (7). On February 28, WHO increased the level of risk of spread and impact of COVID-19 on the global level to very high and declared COVID-19 as a global pandemic on March 11, 2020. (8) However, there is so far no effective treatment or vaccine against the SARS-CoV-2.

At present, guidelines suggest that the use of lopinavir/ritonavir (LPV/r), interferon (IFN) and chloroquine may help to some extent against COVID-19 in adults.(9-11) Seven recently published systematic or rapid reviews suggested that LPV/r, IFN, chloroquine and hydroxychloroquine (HCQ) can be used as an experimental therapy for COVID-19 in adults as the initial treatment, (12-18) but the effectiveness and safety of other antiviral agents (such as ribavirin [RBV], remdesivir and oseltamivir) is uncertain. Although the course of COVID-19 is usually milder in children than adults, children with undeveloped immune system, such as the youngest case confirmed only 30 hours after birth, are at substantial risk of severe infection. (19) Antiviral therapy against SARS-CoV-2 in children is therefore urgently needed, but so far the evidence and literature on the topic remain limited. (20-21)

The objective of this rapid review is to perform a comprehensive literature search and summarize the current evidence on effectiveness and safety of antiviral agents for children with COVID-19. The findings will provide evidence for the development of guideline and the clinical treatment of children with COVID-19.

## Methods

### Search Strategy

Two researchers (Q Shi and X Wang) searched the following electronic databases from their inception until March 31, 2020: MEDLINE (via PubMed), Embase, Web of Science, the Cochrane library, China Biology Medicine (CBM), China National Knowledge Infrastructure (CNKI), and Wanfang Data.(22) We also searched three clinical trial registry platforms (the WHO Clinical Trials Registry Platform, US National Institutes of Health Trials Register and the International Standard Randomized Controlled Trial Number [ISRCTN] Register), Google Scholar, the official websites of WHO and Centers for Disease Control (CDC), and the preprint platforms BioRxiv, MedRxiv, and SSRN. In addition, we searched the reference lists of the identified systematic reviews for further potential studies. Finally, we contacted experts in the field to identify studies that may have been missed.

The search strategy was also peer reviewed by an external specialist. We systematically searched by combining the MeSH and free words. The keywords and terms in the MEDLINE including “COVID-19”, “SARS-CoV-2”, “Novel coronavirus”, “2019-novel coronavirus”, “2019-nCoV”, “antiviral agents”, “antiviral*”, “ribavirin”, “interferon”, “oseltamivir”, “remdesivir”, “lopinavir”, “ritonavir”, “LPV/r” and their derivatives. The details of the search strategy can be found in the *Supplementary Material 1*.

### Inclusion and exclusion criteria

We primarily searched for studies on children less than 18 years of age diagnosed with COVID-19. We made no restrictions on gender, race, or geographical location or setting. COVID-19 was defined according to the WHO interim guidance. (23) If direct evidence on children was unavailable, we also searched indirect evidence from COVID-19 in adults, or from children or adults infected with severe acute respiratory syndrome coronavirus (SARS-CoV) and Middle East respiratory syndrome coronavirus (MERS-CoV) which have similar gene sequences with SARS-CoV-2.

We included all randomized controlled trials (RCTs), clinical controlled trials (CCTs) and cohort studies that compared the effectiveness and safety of antiviral agents (including but not limited to IFN, oseltamivir, LPV/r, RBV, HCQ and remdesivir) with placebo, or comparing the combination of antiviral agents and symptomatic treatment with symptomatic treatment alone. Studies comparing different types and different administration mode of antiviral agents were also included. In vitro studies, animal experiments and basic researches were excluded. Duplicates, articles written in languages other than English or Chinese, conference abstracts and studies where full text could not be retrieved or data were missing were also excluded.

The primary outcomes were mortality and the risk of serious adverse effects (defined as hemolytic anemia, bradycardia and other side effects on cardiovascular system and drug-induced liver injury). The secondary outcomes included the probability of negative PCR test (defined as the rate of negative PCR of SARS-CoV-2 after discharge from the hospital or after receiving antiviral agents which differed in studies), mean or reduction in the dose of corticosteroids, remission of the main clinical symptoms, risk of Acute Respiratory Distress Syndrome (ARDS), duration of disease (defined as the duration (in days) of total stay from symptom onset to recovery), probability of admission to intensive care unit (ICU) and other adverse reactions. All the reasons for exclusion of ineligible studies were recorded, and the process of study selection was documented using a PRISMA flow diagram.

### Study selection

After eliminating duplicates, two researchers (Q Shi and X Wang) independently screened first the titles and abstracts, and then the full-texts of potentially relevant articles, using pre-defined criteria. The specific bibliographic software EndNote was used, and discrepancies were discussed, or solved with a third researcher (Q Zhou). The reasons for exclusion of ineligible studies were recorded. The process of study selection was documented using a PRISMA flow diagram. (23)

### Data extraction

Four researchers (Q Shi, X Wang, Q Zhou and J Liao) extracted data independently in pairs with a pre-determined form, and disagreements were resolved by consensus. We extracted the following data: 1) basic information; 2) participants: baseline characteristics and inclusion/exclusion criteria; 3) details of the intervention and control strategies; and 4) outcomes (for dichotomous data, the number of events and total participants in per group; for continuous data, means, standard deviations (SD), and the number of total participants in per group).

### Risk of bias assessment

Two researchers (Y Yu and Z Wang) independently assessed the potential bias in each included study. Discrepancies were resolved by discussion and consensus to a third researcher (S Lu). For RCTs we used the Cochrane Risk-of-Bias (RoB) assessment tool consisting seven domains: random sequence generation, allocation concealment, blinding of participants and personnel, blinding of outcome assessment, incomplete outcome data, selective outcome reporting, and other bias. (24) We graded each potential source of bias as “Low”, “Unclear” or “High”. For included CCTs, we used ROBINS-I tool, (25) which consists of seven domains: bias due to confounding, bias in selection of participants, bias in classification of interventions, bias due to departures from intended interventions, bias due to missing data, bias in outcome measurement, and bias in selective reporting. The risk of each type of bias was graded as “Low”, “Moderate”, “Serious”, “Critical”, and “No information”. For both RoB and ROBINS-I, the overall risk of bias within each study was based on the results of all the individual domains. For cohort studies, we used Newcastle-Ottawa Scale (NOS) consisting of three domains (selection of exposure, comparability and assessment of outcome). (26) The maximum score was nine, and scores of seven or more was graded as high quality while less than seven scores as low quality.

### Data synthesis

We performed Meta-analyses of outcomes for which the data that were sufficiently compatible. For outcomes with too heterogeneous data, a qualitative synthesis was done. We processed the data according to the Cochrane Handbook by using a random-effects model. (27) For dichotomous data, we calculated risk ratios (RR) with 95% confidence intervals (CI); for continuous data, we calculated weighted mean difference (WMD) with 95% CI. Two-sided *P* values *<* 0.05 were considered statistically significant. (28) Analyses were performed by Stata 14 software (Stata Corp LLC).

For missing SDs, standard errors (SE) were converted to SDs when SE was presented, and if both were missing, we estimated SDs from *P* values or 95% CI. For missing means, we estimated them from interquartile ranges and medians. (29) Statistical heterogeneity was assessed using the chi-square and the *I*^*2*^ statistic, with *P* < 0.10 was considered to be consistent with statistically significant heterogeneity and *I*^*2*^ statistic > 50% indicating substantial heterogeneity. (28) If we detected heterogeneity, we performed subgroup analyses (route, dose, frequency or administration of antivirals) or sensitivity analyses (excluding studies with low-quality or high risk of bias; excluding studies in which mean or SD, or both of them were imputed for missing data) to explore the reasons. Publication bias was assessed by examining the symmetry of the funnel-plot.

### Quality of the evidence assessment

We assessed the quality of evidence using the Grading of Recommendations Assessment, Development and Evaluation (GRADE) approach, and classified the evidence quality as “high”, “moderate”, “low” and “very low”. (30-31) We also produced “Summary of Findings”tables. Direct evidence from RCTs starts at high quality, and evidence from observational studies at low quality. In the next step, the quality can be downgraded for five different reasons (study limitations, consistency of effect, imprecision, indirectness, and publication bias) and upgraded for three reasons (large magnitude of effect, dose-response relation and plausible confounders or biases).

## Results

### Study and Patient Characteristics

We identified 4095 references from the databases, and six records from additional searches. A total of 1216 records were excluded as duplicates, after screening for titles, abstract and full texts, no direct evidence for children with COVID-19 was found. Finally, a total of 23 studies (six RCTs and 17 cohort studies) with 6008 patients of indirect evidence were included (*Figure 1*) (32-54). These studies were published between 2003 and 2020 and the sample size ranged from 22 to 1701, of which, seven studies were on COVID-19, 13 studies on SARS and three studies on MERS. Another study of Cai 2020 was found but in temporary removal condition, therefore it was not included (55).

**Figure 1.**
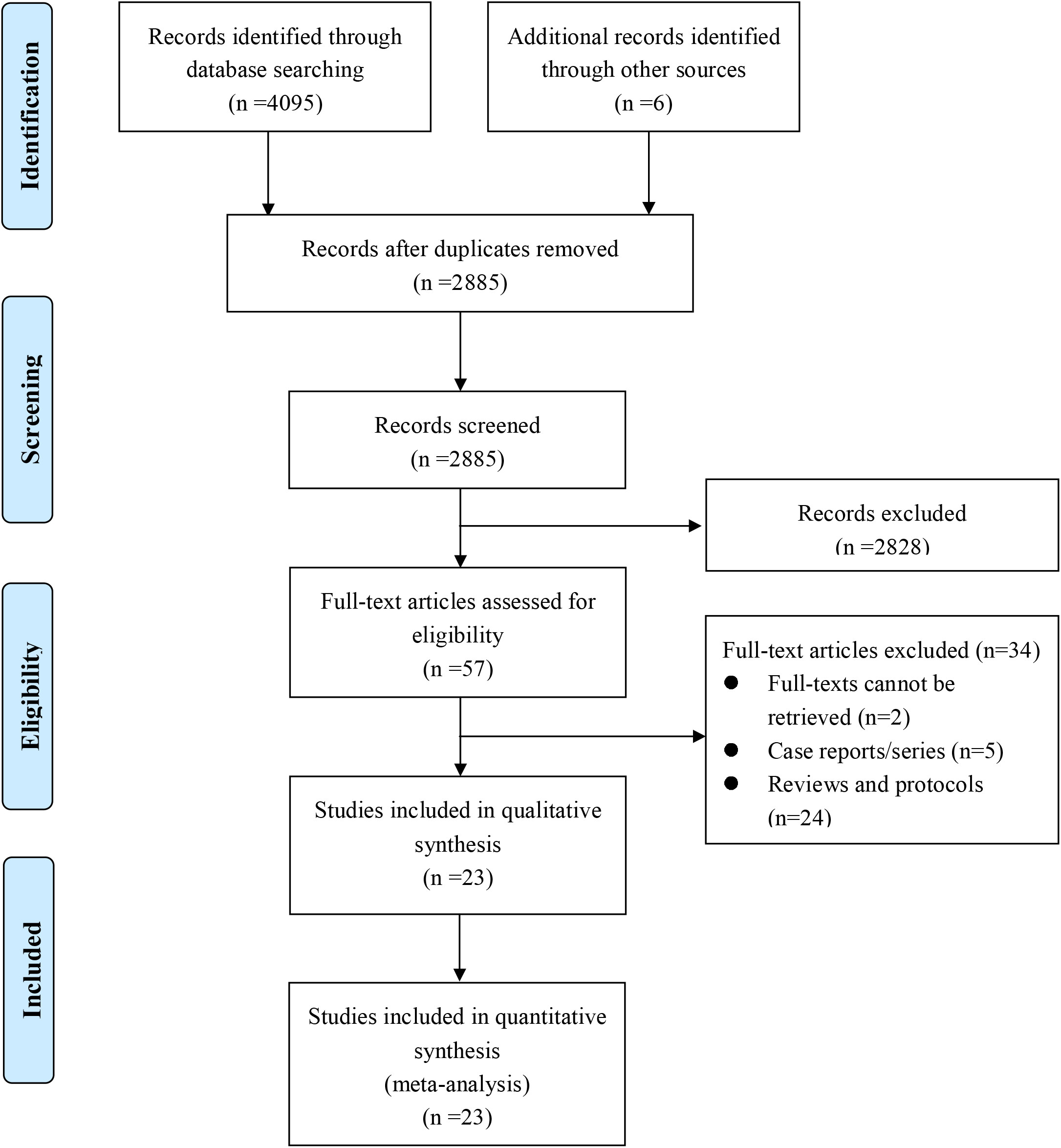
Flow diagram of the literature search. Abbreviations: CBM: China Biology Medicine; CNKI: China National Knowledge Infrastructure; WHO: World Health Organization; CDC: Centers for Disease Control; COVID-19: Corona Virus Disease hyphen one nine; SARS: Severe Acute Respiratory Syndrome; MERS: Middle East respiratory syndrome; ARDS: Acute Respiratory Distress Syndrome.

The risk of bias in the included three RCTs were high, as they did not perform allocation concealment and blindness for patients and clinicians. The other three RCTs had low risk of bias (n=3). More than half of the cohort studies (n=9) had a high risk of bias, the main reasons were being the lack of controlling for important factors that would influence the primary study results, lack of long enough follow-up for outcomes to occur, and inadequate outcome ascertainment. Study characteristics and risk of bias are illustrated in *Table 1*.

**Table 1.** Baseline Characteristics of 23 Included Studies

| Study | Country | Design | Sample | Disease | Sample size |  | Age (year)<sup>†</sup> |  | Sex (Male/Female) |  | Risk of bias |
| --- | --- | --- | --- | --- | --- | --- | --- | --- | --- | --- | --- |
|  |  |  |  |  | Intervention | Control | Intervention | Control | Intervention | Control |  |
| Cao 2020 (33) | China | RCT | 199 | COVID-19 | 99 | 100 | 58.7 ± 13.5 | 58.0 ± 15.0 | 61/38 | 59/41 | High |
| Chen 2020 (53) | China | RCT | 236 | COVID-19 | 116 | 120 | NR | NR | 59/57 | 51/69 | High |
| Li 2020 (34) | China | RCT | 44 | COVID-19 | 37 | 7 | 49.4 ± 14.9 |  | 21/23 |  | Low |
| Chen 2020 (54) | China | RCT | 30 | COVID-19 | 15 | 15 | 50.5 ± 3.8 | 46.7 ± 3.6 | 9/7 | 12/15 | High |
| Chen 2020 (55) | China | RCT | 62 | COVID-19 | 31 | 31 | 44.1 ± 16.1 | 45.2 ± 14.7 | 14/17 | 15/16 | Low |
| Xu 2011 (50) | China | RCT | 127 | SARS | 77 | 50 | 44.4 ± 16.3 | 34.8 ± 12.8 | 59/68 |  | Unclear |
| Chen 2020 (35) | China | Cohort | 134 | COVID-19 | 86 | 48 | 48 (35-62) |  | 69/65 |  | 5 |
| Deng 2020 (38) | China | Cohort | 33 | COVID-19 | 16 | 17 | 41.8 ± 14.08 | 47.25 ± 17.25 | 7/9 | 10/7 | 7 |
| Chan 2003 (36) | China | Cohort | 678 | SARS | 44 | 634 | NR | NR | 12/32 | NR | 7 |
| Chu 2003 (37) | China | Cohort | 152 | SARS | 41 | 111 | 39.4 ± 15.2 | 42.1 ± 14.7 | 10/31 | 48/63 | 6 |
| Loutfy 2003 (39) | Canada | Cohort | 22 | SARS | 9 | 13 | 44.8 ± 9.7 | 46.5 ± 20.9 | 3/6 | 3/10 | 7 |
| Leong 2004 (44) | Singapore | Cohort | 229 | SARS | 97 | 132 | 34.4 ± 14.3 | 42.6 ± 17.7 | 22/75 | 51/81 | 7 |
| Guo 2019 (49) | China | Cohort | 103 | SARS | 34 | 69 | 29.9 ± 10.1 | 37.0 ± 13.2 | 11/23 | 33/36 | 7 |
| Muller 2007 (48) | Canada | Cohort | 306 | SARS | 183 | 123 | 44 (34–56) | 45 (36–57) | 73/110 | 41/82 | 7 |
| Chiou 2005 (45) | China | Cohort | 51 | SARS | 44 | 7 | 36.4 ± 15.7 | 49.8 ± 26.1 | 11/33 | 2/5 | 6 |
| Lau 2009 (46) | China | Cohort | 1104 | SARS | 309 | 795 | NR | NR | 125/184 | 395/400 | 6 |
| Liu 2009 (41) | China | Cohort | 1701 | SARS | 1200 | 501 | 42.3 ± 14.8 |  | 801/900 |  | 5 |
| Wang 2005 (47) | China | Cohort | 90 | SARS | 53 | 37 | 36.7 ± 13.7 | 39.6 ± 16.0 | 60/30 |  | 5 |
| Li 2005 (40) | China | Cohort | 87 | SARS | 41 | 46 | 29.3 ± 10.6 | 26.7 ± 8.2 | 8/33 | 8/38 | 7 |
| Xu 2003 (42) | China | Cohort | 203 | SARS | 83 | 120 | 41.1 ± 17.7 |  | 123/138 |  | 5 |
| Omrani 2014 (51) | Saudi Arabia | Cohort | 44 | MERS | 20 | 24 | 67.4 ± 18.5 | 64.0 ± 18.1 | 16/4 | 16/8 | 8 |
| Arabi 2019 (52) | Saudi Arabia | Cohort | 349 | MERS | 144 | 205 | 58 (47-70) | 58.0 (41-70) | 101/43 | 140/65 | 6 |
| Shalhoub 2015 (43) | Saudi Arabia | Cohort | 24 | MERS | 13 | 11 | 65 (33–84) | 67 (25–88) | 10/3 | 4/7 | 6 |

**Abbreviations:** NR: Not Reported; RCT: Randomized Controlled Trial; COVID-19: Corona Virus Disease hyphen one nine; SARS: Severe Acute Respiratory Syndrome; MERS: Middle East respiratory syndrome.
<sup>†</sup>Ages were reported either as mean ± standard deviation, or median (interquartile range); Sex (Male/Female) was reported as number

### Efficacy and Safety of Existing Antiviral Agents

The results of the Meta-analysis for each type of antiviral agent are shown in *Table 2*. The details of primary data from each retrieved study can be found in *Supplementary Material 2*. The details of GRADE for each outcome can be found in *Supplementary Material 3*. Due to the small number of studies for each outcome, we were unable to evaluate publication bias.

**Table 2.**
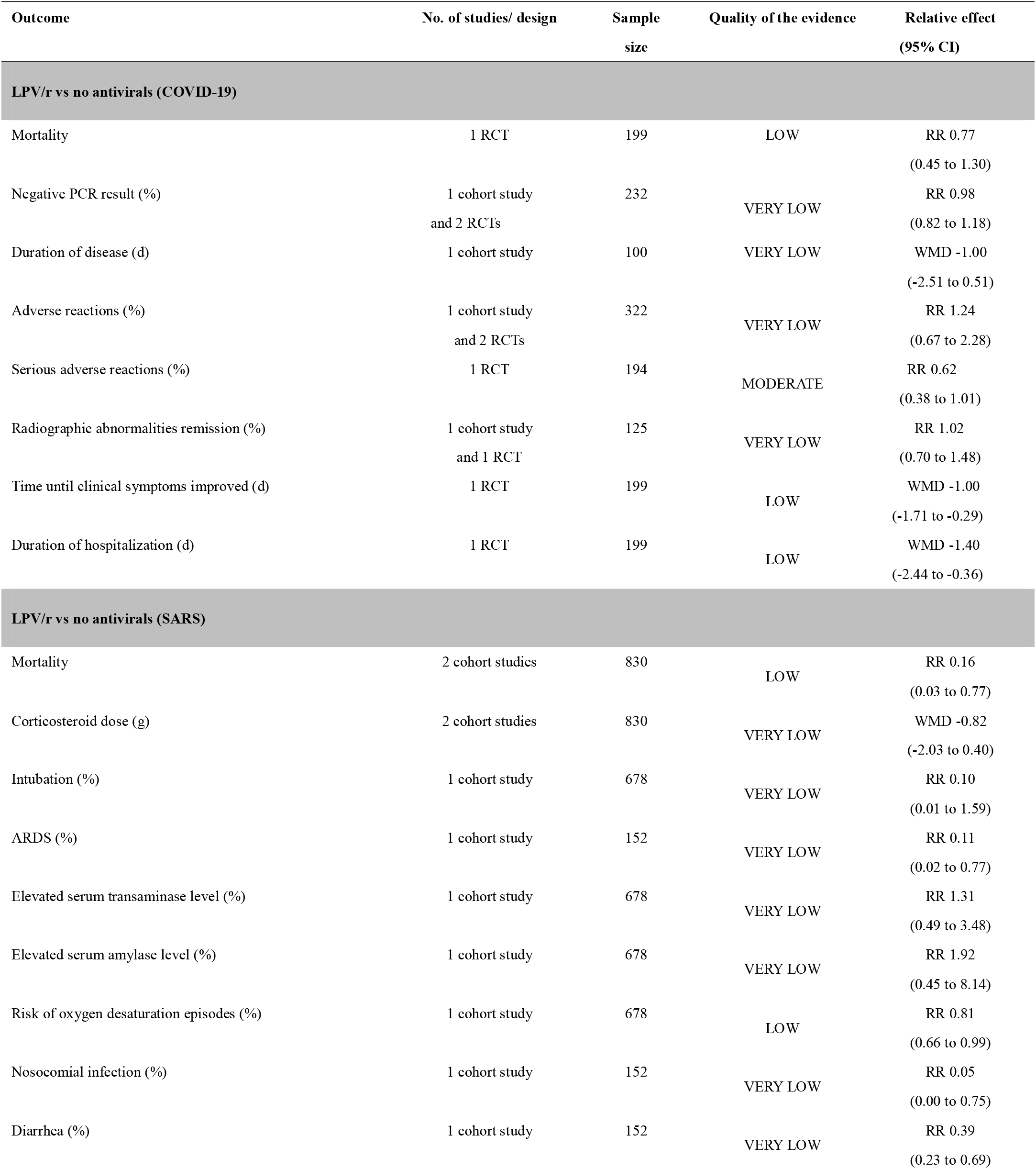
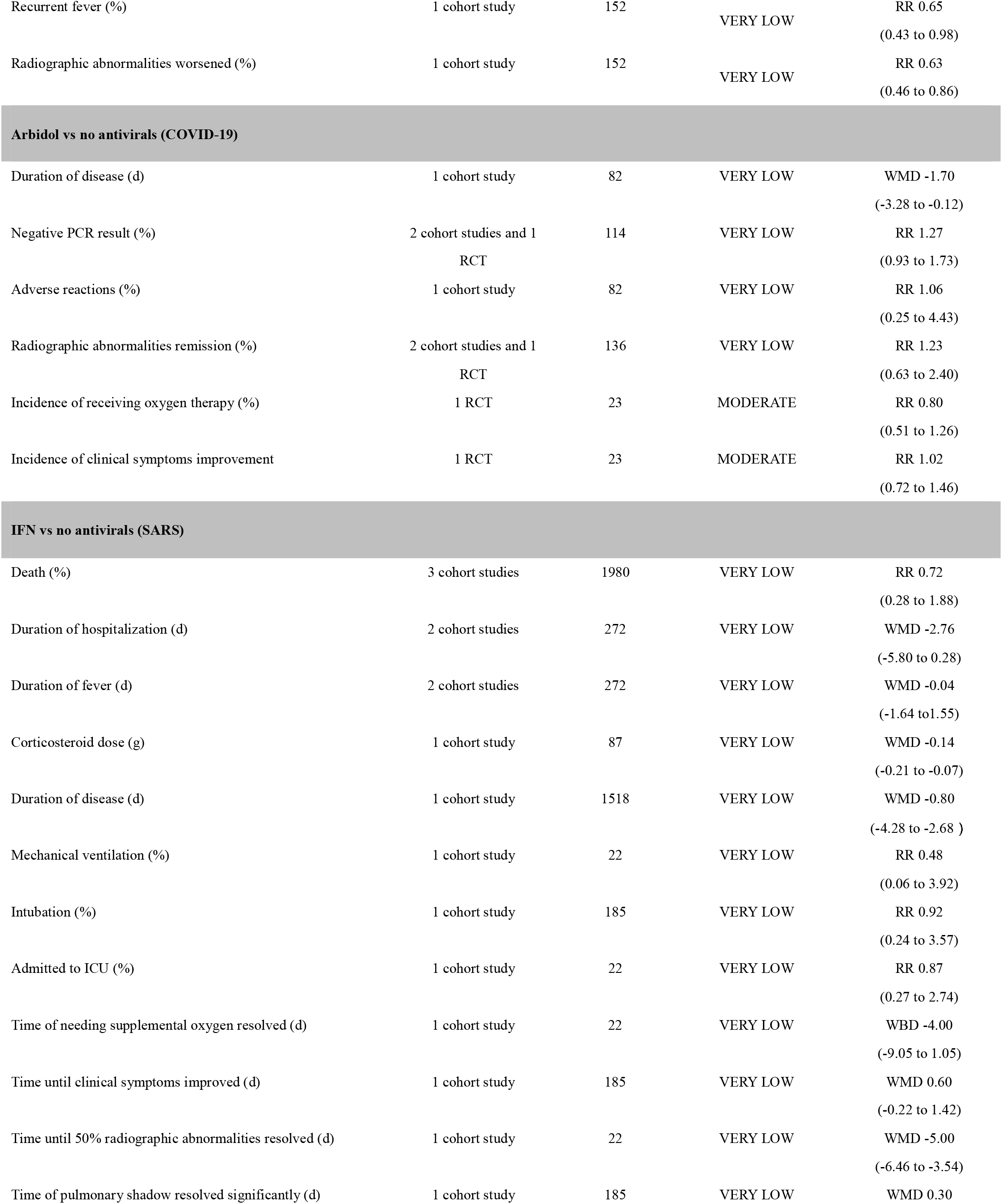
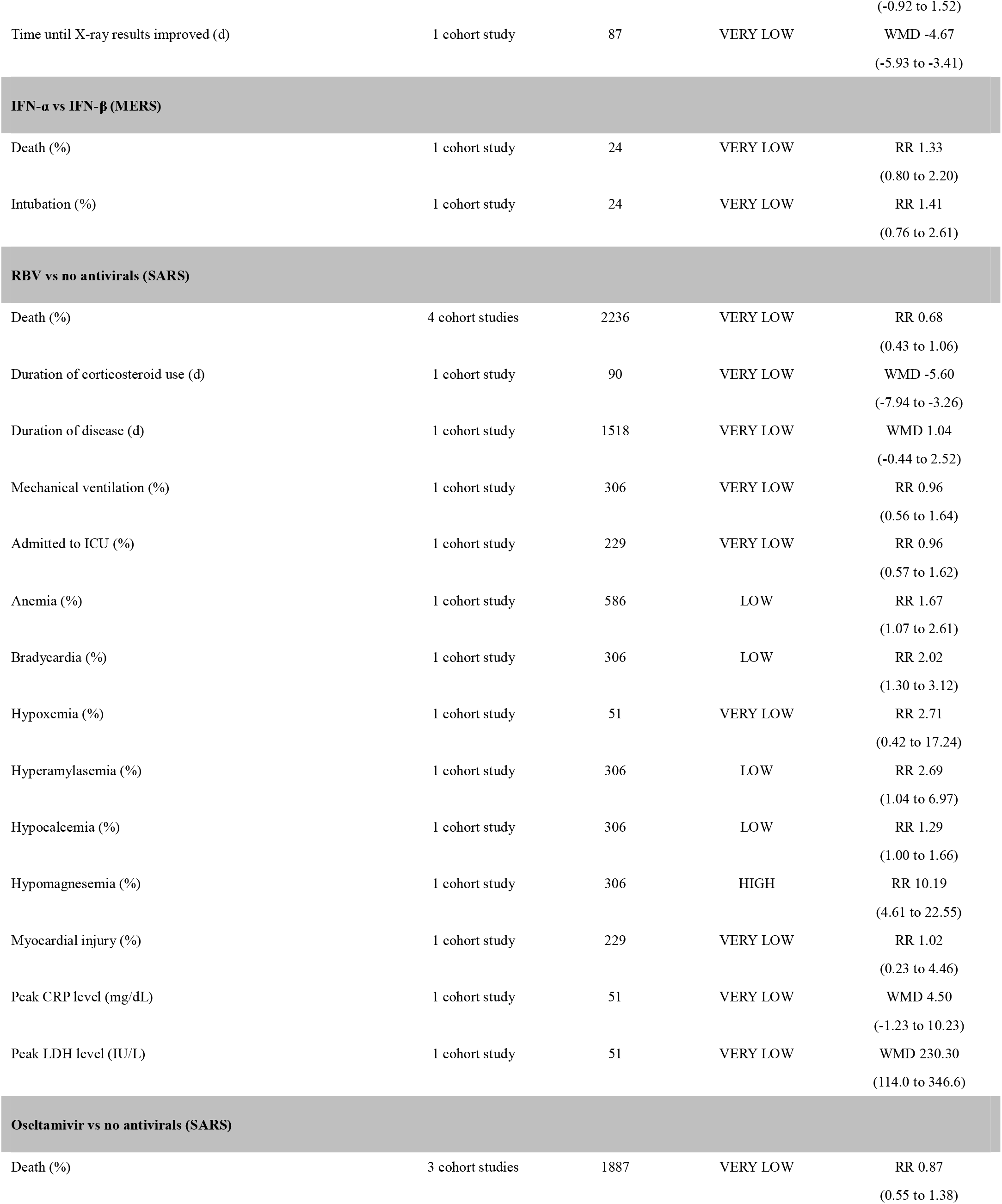
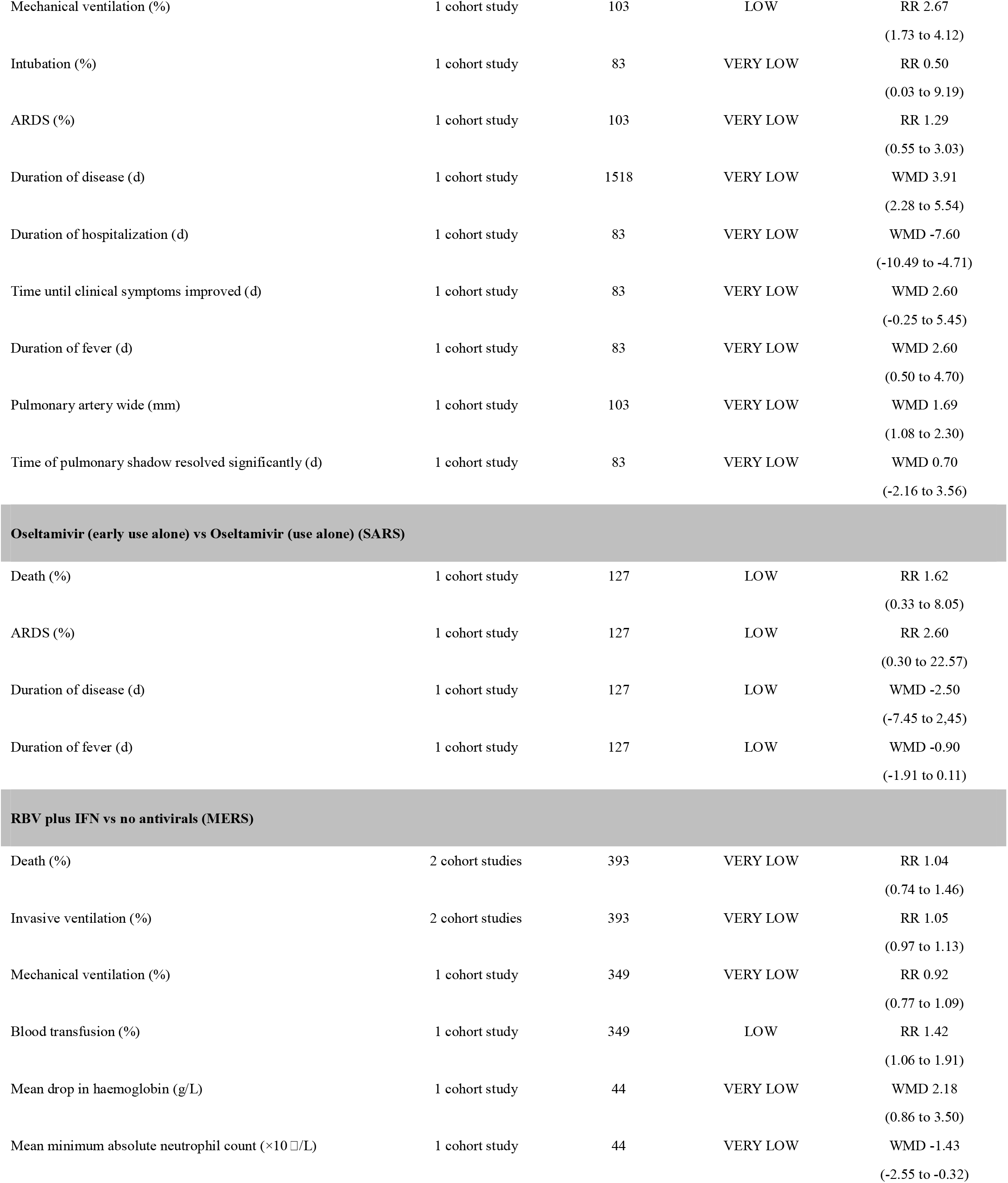
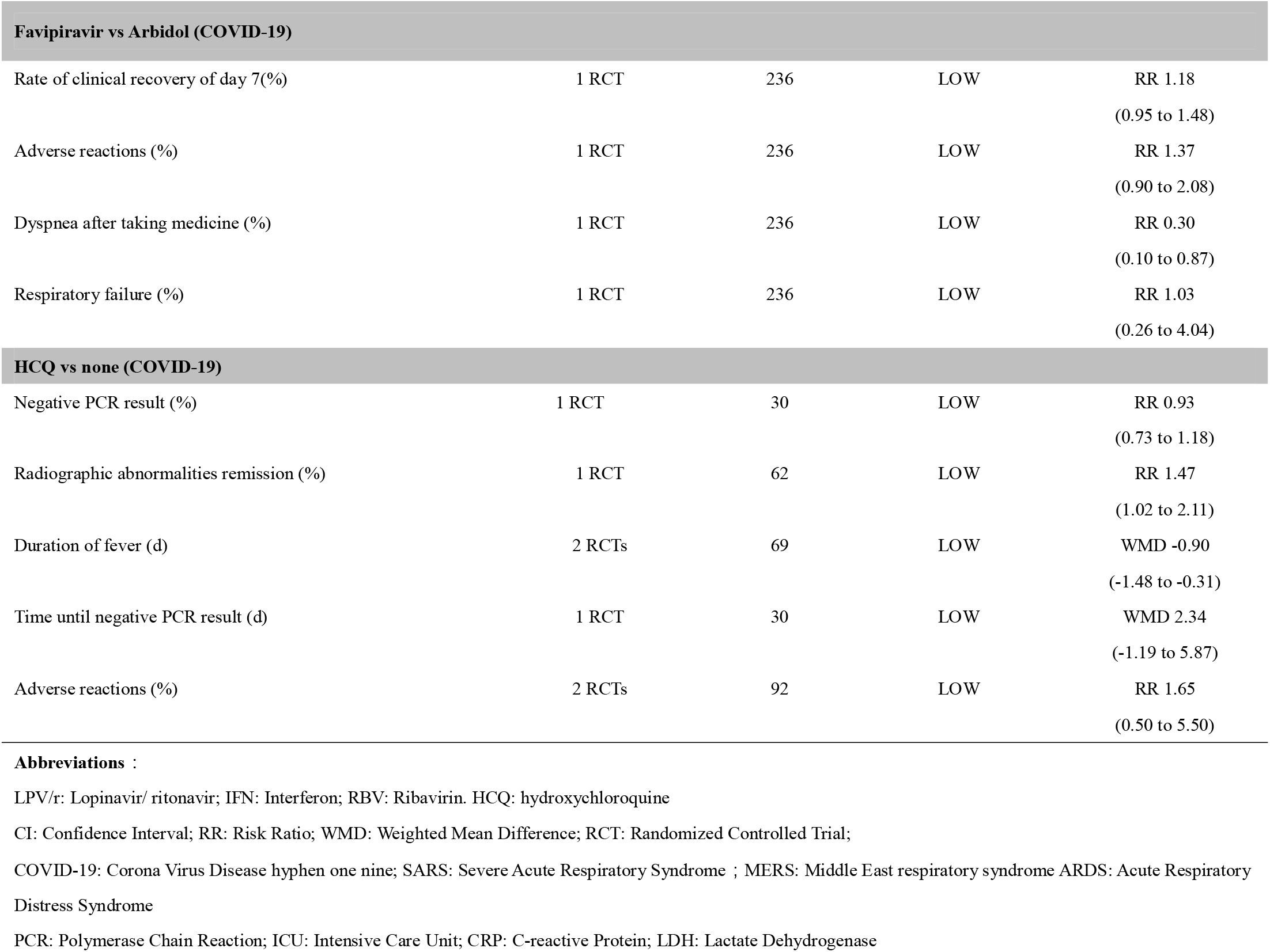
Summary of evidence for the effectiveness and safety of antiviral agents

### Lopinavir/ritonavir

Three studies with a total of 327 patients (32-34) reported the effectiveness and safety of LPV/r in adult patients with COVID-19. There was no statistically significant difference in the mortality (RR =0.77, 95% CI 0.45 to 1.30, low-quality evidence, *Figure 2*) and probability of negative PCR test (RR=0.98, 95 CI% 0.82 to 1.18, very low-quality evidence) between the intervention and control groups. There was also no statistically significant difference in the incidence of adverse reactions (RR=1.24, 95 CI% 0.67 to 2.28, very low-quality evidence) and serious adverse reactions (RR=0.62, 95 CI% 0.38 to 1.01, moderate-quality evidence) between the two groups, of which, the most common side effects were gastrointestinal reaction (including nausea and vomiting, diarrhea and abnormal liver function).

**Figure 2.**
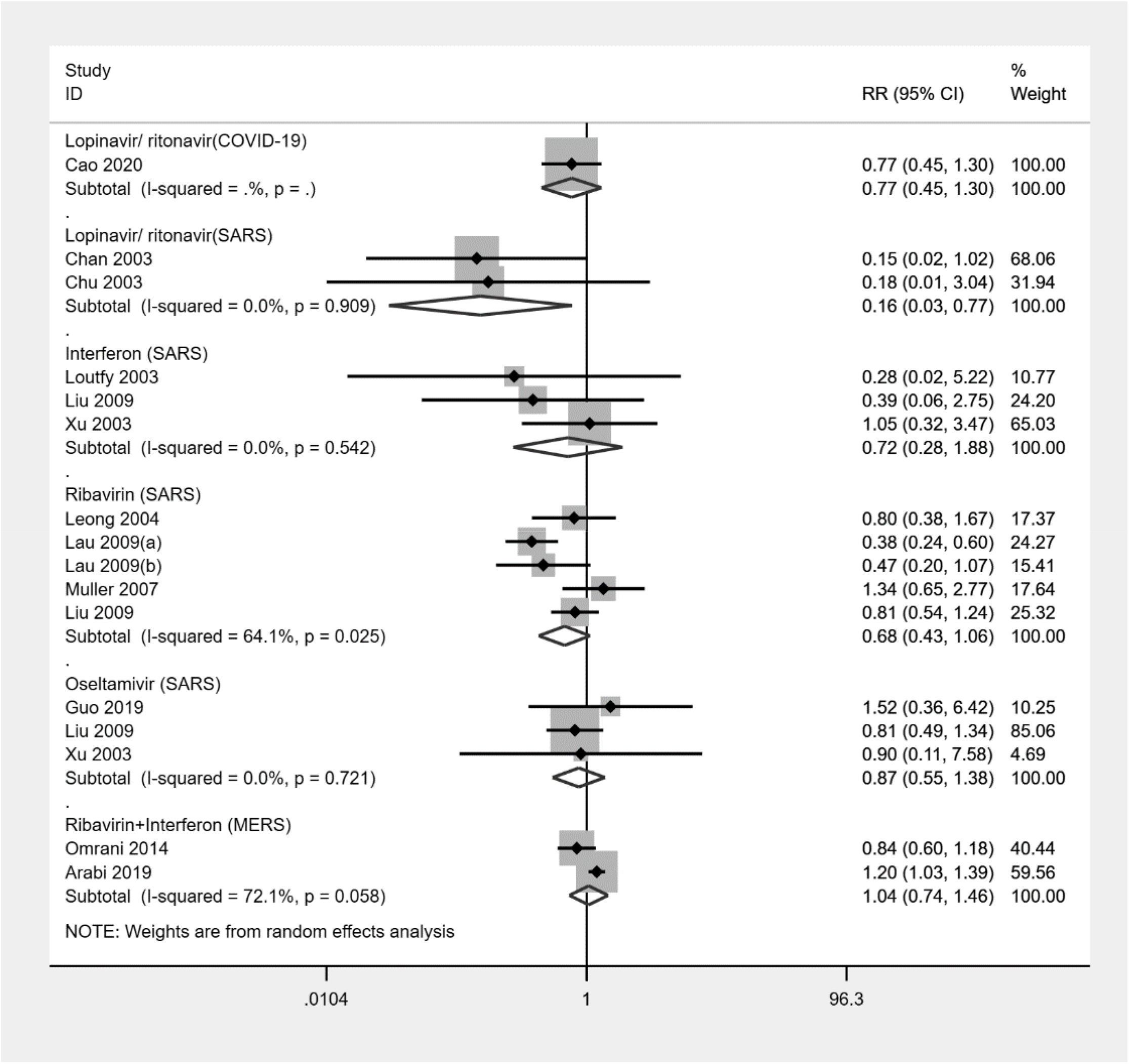
Forest plot of mortality for included studies comparing antivirals with no antivirals. Abbreviations: COVID-19: Corona Virus Disease hyphen one nine; SARS: Severe Acute Respiratory Syndrome;MERS: Middle East respiratory syndrome. The results of Meta-analysis indicated that lopinavir/ritonavir had no effect on mortality in adults with COVID-19 (risk ratio [RR]= 0.77, 95% confidence interval [CI] 0.45 to 1.30), but could decrease the mortality in adults with SARS (RR=0.16, 95% CI 0.03 to 0.77), and interferon (RR=0.72, 95% CI 0.28 to 1.88), ribavirin (RR=0.68, 95% CI % 0.43 to 1.06), oseltamivir (RR=0.87, 95% CI 0.55 to 1.38) did not reduce the mortality in adults with SARS, while combination of ribavirin and interferon was not efficeive for reducing the mortality in adults with MERS (RR=1.04, 95% CI 0.74 to 1.46).

Two cohort studies with a total of 830 patients (35,36) reported the effectiveness and safety of LPV/r in adult patients of SARS. The results showed that LPV/r therapy decreased the risk of death (RR=0.16, 95% CI 0.03 to 0.77, low-quality evidence, *Figure 2*) and ARDS (RR=0.11, 95% CI 0.02 to 0.77, very low-quality evidence) compared with the control group. However, no statistically significant difference was found in the dose of corticosteroids (WMD=-0.82 g, 95% CI -2.03 to 0.40) with considerable heterogeneity of the *I*-squared was 86.4%, because both means and SDs of the two studies were imputed from missing data. In addition, patients in the LPV/r group were more often nosocomially infected (RR=0.05, 95% CI 0.00 to 0.75), and had a higher risk of adverse reactions such as diarrhea (RR=0.39, 95% CI 0.23 to 0.69) or recurrent fever (RR=0.65, 95% CI 0.43 to 0.98). The overall quality of evidence was very low.

### Arbidol

Three studies with a total of 138 patients (33-34,37) reported the effectiveness and safety of arbidol in adult patients of COVID-19. There was no statistically significant difference in the probability of having a negative PCR result (RR=1.27, 95% CI 0.93 to 1.73), probability of radiographic abnormalities remission (RR=1.23, 95% CI 0.63 to 2.40) and duration of disease (WMD=-1.70 days, 95% CI -3.28 to -0.12) between patients with arbidol therapy and control group. Because of the large heterogeneity in the radiographic abnormalities remission, we performed a subgroup analysis of study design, and we still found no significant association in neither cohort studies (RR=1.58, 95% CI 0.97 to 2.59) nor RCTs (RR=0.71, 95% CI 0.47 to 1.06). There was also no statistically significant difference in the incidence of adverse reactions (RR=1.06, 95% CI 0.25 to 4.43) between the two groups. The overall quality of evidence was very low.

### Interferon

Four cohort studies with a total of 2013 patients (38-41) reported the effectiveness and safety of intramuscular or subcutaneous injection of IFN in adult patients with SARS. The results showed that IFN therapy decreased the dose of corticosteroids dose (WMD=-0.14 g, 95% CI -0.21 to -0.07) and promoted the remission of radiographic abnormalities. No statistically significant difference was found in mortality (RR=0.72, 95% CI 0.28 to 1.88, *Figure 2*). No obvious adverse reactions were reported in any of the above four studies. The quality of evidence was very low.

One cohort study with a total of 24 patients (43) compared the effectiveness of different types of IFN in adult patients with SARS. The results showed that there was no statistically significant difference in the risk of death (RR=1.33, 95% CI 0.80 to 2.20) between patients treated with IFN-α and IFN-β. The quality of evidence was very low.

### Ribavirin

Six cohort studies with a total of 3481patients (40,43-47) reported the effectiveness and safety of RBV in adult patients with SARS. The results showed that RBV therapy significantly decreased the duration of corticosteroid use (WMD =-5.60 g, 95% CI -7.94 to -3.26, very low-quality evidence), and increased the duration of disease (WMD=1.04 d, 95% CI -0.44 to 2.52, very low-quality evidence) compared with the control group. There was no statistically difference in the risk of death (RR=0.68, 95% CI % 0.43 to 1.06, *Figure 2*). In addition, the use of RBV was associated with an increased risk of adverse reactions, including anemia (RR=1.67, 95% CI 1.07 to 2.61, low-quality evidence), bradycardia (RR=2.02, 95% CI 1.30 to 3.12, low-quality evidence), and hypomagnesemia (RR=10.19, 95% CI 4.61 to 22.55, high-quality evidence).

### Oseltamivir

Three cohort studies with a total of 2007 patients (40-41,48) reported the effectiveness and safety of oseltamivir in adult patients with SARS. The results showed that there was no statistically significant difference in the risk of death (RR=0.87, 95% CI 0.55 to 1.38, *Figure 2*) between oseltamivir therapy and the control group. The use of oseltamivir prolonged the duration of disease (WMD=3.91d, 95% CI 2.28 to 5.54, very low-quality evidence) and duration of fever (WMD=2.60 d, 95% CI 0.50 to 4.70, very low-quality evidence).

One RCT with a total of 127 patients (49) compared the effectiveness of oseltamivir between early use alone and use alone in adult patients with SARS. The results showed that early use alone was not associated with the risk of death (RR=1.62, 95% CI 0.33 to 8.05), ARDS (RR=2.60, 95% CI 0.30 to 22.57) or the duration of disease (WMD=-2.50 d, 95% CI -7.45 to 2.45). The quality of evidence was low.

### Combination of ribavirin and interferon

Two cohort studies with a total of 393 patients (50-51) reported the effectiveness and safety of a combination of RBV and IFN for adult patients with MERS. The results showed that combination therapy of RBV and IFN could increase the mean reduction in hemoglobin (WMD=2.18 g/L, 95% CI 0.86 to 3.50, very low-quality evidence) and the need of blood transfusion (RR=1.42, 95% CI 1.06 to 1.91, low-quality evidence). But there was no statistically significant difference in the risk of death (RR=1.04, 95% CI 0.74 to 1.46, *Figure 2*) between the two groups.

### Favipiravir

One study with a total of 236 patients (52) reported the effectiveness and safety of favipiravir for adult patients with COVID-19. The results showed that when comparing to arbidol, favipiravir had lower incidence of dyspnea after taking medicine (RR=0.30, 95% CI 0.10 to 0.87), but there was no differnce in clinical recovery (RR=1.18, 95% CI 0.95 to 1.48) or the incidence of adverse reactions (RR=1.37, 95% CI 0.90 to 2.08). The overall quality of evidence was low.

### Hydroxychloroquine

Two studies with a total of 92 patients (53-54) reported the effectiveness and safety of hydroxychloroquine for adult patients with COVID-19. The results showed that HCQ had no benefit on the negative PCR result (RR=0.93, 95% CI 0.73 to 1.18), but was effective for shortening the duration of fever (WMD=-0.90 days, 95% CI -1.48 to -0.31). In addition, there was no statistically significant difference in the incidence of adverse reactions (RR=1.65, 95% CI 0.50 to 5.50) between HCQ therapy and the control group. The overall quality of evidence was low.

## Discussion

Our rapid review identified a total of 23 studies. No direct evidence for the effectiveness and safety of antiviral agents for children with COVID-19 was available. Based on the analysis of indirect evidence from adult patients with COVID-19, very low to low-quality evidence indicated that LPV/r, arbidol and hydroxychloroquine were not effective. For adult patients with SARS or MERS, very low to low-quality evidence indicated that LPV/r, IFN, RBV and oseltamivir had no clinical effectiveness on mortality, corticosteroids dose, or other main outcomes. Certain medications, such as LPV/r and RBV, were likely to lead to adverse reactions (such as gastrointestinal reaction, abnormal liver function, anemia, bradycardia, or hypoxemia).

Most viral diseases are self-limiting illnesses that do not require specific antiviral therapy. At present, no antiviral agent has been confirmed to be effective against COVID-19, and vaccination are currently under development, so symptomatic and supportive treatments are crucial. However, children are less likely than adults to have complications or develop into critical conditions, and their clinical manifestations are less atypical, complicating the diagnosis. (56-58) Guidelines recommend antiviral agents such as LPV/r, IFN, arbidol and chloroquine to treat COVID-19 in adults, while children (especially critically illness) can be treated reference to the regimen of adults. (9-11) Up to now, almost all COVID-19 patients (adults and children) have received antiviral therapy.(59) Several case reports or series (60-61) have also highlighted the potential efficacy of antivirals in children with SARS-CoV-2 infection and found no obvious adverse reactions, but the number was too small to draw any conclusions. More studies are needed to further evaluate the risks and benefits that antiviral agents may bring.

LPV/r is one of the first medications that were taken into clinical practice after the beginning of the COVID-19 outbreak, and it is recommended for treatment of COVID-19 patients in the latest version of China national practice guideline (released on March 4, 2020) without any reference. (9) Our rapid review however demonstrates that LPV/r is unlikely to be effective for COVID-19 in adults with numerous obvious adverse reactions, which was the same as recent studies. (62-63) Two rapid reviews conducted in 2020 (12-13) examined that early use of LPV/r can reduce the mortality and steroid dosing in patients with SARS and MERS, and suggested that it could be used as a component for an experimental regimen to treat COVID-19. But no quantitative analysis or evidence grading was performed, and therefore the reliability of the conclusions is questionable. Although LPV/r could reduce the mortality of adult SARS patients, the quality of evidence was low. Therefore, LPV/r should not be recommended in clinical practice guidelines.

The results on other antivirals were similar to those identified in other systematic reviews. Among patients with COVID-19, the use of HCQ was effective for clinical recovery which is the same as published reviews (17-18), but we found HCQ had no benefit on probability of viral load disappearance, this is not the same as previous studies due to the retraction of Philippe 2020 (64). All trials for HCQ included in this study had small sample size to draw robust conclusions. IFN had no benefit on mortality and the effect did not differ between IFN-α and IFN-β: the results are in line with another recent rapid review. (14) RBV and oseltamivir were not shown effective for treating adults with SARS, and the use of RBV was even related to a high risk of serious side effects, and oseltamivir prolonged the duration of disease. These results were also observed by recent and previous systematic reviews. (15, 65-66) One study demonstrated that the concentration of RBV required to effectively inhibit the activity of SARS-CoV or MERS-CoV was beyond the clinically acceptable range, so routine use of the drug would have no effect. (67) One recent case of COVID-19 in the United States suggested a promising clinical response to remdesivir, (68) and study by Wang et al. revealed that remdesivir was highly effective in the control of SARS-CoV-2 in vitro, (69) but the evidence quality was low and the newest results of clinical research suggested no significant effect for patients hospitalized of severe COVID-19, (70) and the clinical trials of remdesivir therapy are still ongoing. The outbreak of COVID-19 has imposed a great socioeconomic, public health, and clinical burden on the affected countries and regions, especially for the low-and middle-income countries. Therefore, priority should be given for the research and implementation of agents with promising outcomes.

### Strengthens and Limitations

This study is to our knowledge the first systematic and comprehensive rapid review for the effectiveness and safety of antiviral agents in children with COVID-19. It can therefore be considered the best evidence at the moment for the management of COVID-19 in children, and help to respond to the current public health emergency. Our study was also performed and reported in accordance with Cochrane Handbook and PRISMA checklist, and included Meta-analyses and grading of evidence to draw quantitative conclusions with scientific and rigorous methods. However, our study had also some limitations: First, this rapid review was unable to identify direct evidence for antiviral use in children with COVID-19 and only summarized the indirect evidence, mainly from adults patients with COVID-19, SARS or MERS. The reported treatment effects should be interpreted with caution due to the lack of high-quality RCTs and direct evidence. Second, due to the heterogeneity of the reviewed studies in terms of the wide range of treatment dosages, frequencies and routes of administration, we were unable to perform a quantitative analysis from these aspects for each antiviral. This is a major obstacle to a clear interpretation of the results of this review. Third, because of the specificity and urgency of PHEIC, our study protocol was not registered on the Prospective Register of Systematic Reviews Platform.

### Further Suggestions

We suggest for the further actions on the basis of our study. First, high-quality clinical research should be carried out in a timely and effective manner, following the randomization, control and bind principles of evidence-based medicine, trying to adopt objective and representative outcomes for evaluation, so that unbiased research results can be ensured. Second, health workers need high-quality, unbiased and evidence-based recommendations to guide clinical practice. Health workers should accumulate clinical experience and be encouraged to interpret the evidence with professionalism by cooperating with researchers, avoid conflicts of interest, and thus reduce the possibly harmful impact on children with COVID-19. Third, health policy decisions should be made based on the best available evidence and make full use of the limited resources to make decisions that are valid, rational and based on up-to-date scientific knowledge.

### Conclusions

In conclusion, there is no direct evidence for antiviral agents in children with COVID-19 so far. Very low to low-quality indirect evidence indicated that antiviral agents were not effective for reducing mortality, and the effectiveness and safety of antivirals for children with COVID-19 are uncertain. Therefore, we cannot suggest routine use of these agents for the treatment of COVID-19 in children, with the exception of clinical trials after thorough ethical assessment.

## Data Availability

The data to support the findings of this study was extracted from the included studies, and the reliability was guaranteed.

## Author contributions

(I) All the authors contributed to the conception and design of the study as follows; (II) Administrative support: Y Chen; (III) Provision of study materials or patients: Q Shi, Q Zhou and X Wang; (IV) Collection and assembly of data: Q Shi, Q Zhou, X Wang, J Liao; (V) Data analysis and interpretation: Q Shi, Q Zhou and X Wang, J Liao, Y Yu, Z Wang, S Lu; (VI) Manuscript writing: All authors; (VII) Final approval of manuscript: All authors.

## Acknowledgments

We thank Janne Estill, Institute of Global Health of University of Geneva for providing guidance and comments for our review. We thank all the authors for their wonderful collaboration

## Ethical Statement

The authors are accountable for all aspects of the work in ensuring that questions related to the accuracy or integrity of any part of the work are appropriately investigated and resolved.

## Supplementary Material 1-Search strategy PubMed

#1 “COVID-19” [Supplementary Concept]

#2 “Severe Acute Respiratory Syndrome Coronavirus” [Supplementary Concept] #3 “Middle East Respiratory Syndrome Coronavirus” [Mesh]

#4 “Severe Acute Respiratory Syndrome” [Mesh]

#5 “SARS Virus” [Mesh]

#6 “COVID-19” [Title/Abstract]

#7 “SARS-COV-2” [Title/Abstract]

#8 “Novel coronavirus” [Title/Abstract]

#9 “2019-novel coronavirus” [Title/Abstract]

#10 “coronavirus disease-19” [Title/Abstract]

#11 “coronavirus disease 2019” [Title/Abstract]

#12 “COVID19” [Title/Abstract]

#13 “Novel CoV” [Title/Abstract]

#14 “2019-nCoV” [Title/Abstract]

#15 “2019-CoV” [Title/Abstract]

#16 “Wuhan-Cov” [Title/Abstract]

#17 “Wuhan Coronavirus” [Title/Abstract]

#18 “Wuhan seafood market pneumonia virus” [Title/Abstract]

#19 “Middle East Respiratory Syndrome” [Title/Abstract]

#20 “MERS” [Title/Abstract]

#21 “MERS-CoV” [Title/Abstract]

#22 “Severe Acute Respiratory Syndrome” [Title/Abstract]

#23 “SARS” [Title/Abstract]

#24 “SARS-CoV” [Title/Abstract]

#25 “SARS-Related” [Title/Abstract]

#26 “SARS-Associated” [Title/Abstract]

#27 #1-#26/ OR

#28 “Antiviral Agents” [Mesh]

#29 “Ribavirin” [Mesh]

#30 “Interferon” [Mesh]

#31 “GS-5734” [Supplementary Concept]

#32 “Oseltamivir” [Mesh]

#33 “Lopinavir” [Mesh]

#34 “Ritonavir” [Mesh]

#35 “lopinavir-ritonavir drug combination” [Supplementary Concept]

#36 “Antiviral*” [Title/Abstract]

#37 “Ribavirin” [Title/Abstract]

#38 “Virazole” [Title/Abstract]

#39 “Interferon” [Title/Abstract]

#40 “Remdesivir” [Title/Abstract]

#41 “GS-5734” [Title/Abstract]

#42 “Oseltamivir” [Title/Abstract]

#43 “Lopinavir” [Title/Abstract]

#44 “Ritonavir” [Title/Abstract]

#45 “Kaletra” [Title/Abstract]

#46 “LPV/r” [Title/Abstract]

#47 #28-#46/ OR

#48 #27 AND #47

### Embase

#1 ‘middle east respiratory syndrome coronavirus’/exp

#2 ‘severe acute respiratory syndrome’/exp

#3 ‘sars coronavirus’/exp

#4 ‘COVID-19’:ab,ti

#5 ‘SARS-COV-2’:ab,ti

#6 ‘novel coronavirus’:ab,ti

#7 ‘2019-novel coronavirus’:ab,ti

#8 ‘coronavirus disease-19’:ab,ti

#9 ‘coronavirus disease 2019’:ab,ti

#10 ‘COVID19’:ab,ti

#11 ‘novel cov’:ab,ti

#12 ‘2019-ncov’:ab,ti

#13 ‘2019-cov’:ab,ti

#14 ‘wuhan-cov’:ab,ti

#15 ‘wuhan coronavirus’:ab,ti

#16 ‘wuhan seafood market pneumonia virus’:ab,ti

#17 ‘middle east respiratory syndrome’:ab,ti

#18 ‘middle east respiratory syndrome coronavirus’:ab,ti

#19 ‘mers’:ab,ti

#20 ‘mers-cov’:ab,ti

#21 ‘severe acute respiratory syndrome’:ab,ti

#22 ‘sars’:ab,ti

#23 ‘sars-cov’:ab,ti

#24 ‘sars-related’:ab,ti

#25 ‘sars-associated’:ab,ti

#26 #1-#25/ OR

#27 ‘Antiviral Agent’/exp

#28 Antiviral*[ti, ab]

#29 Ribavirin [ti, ab]

#30 Virazole [ti, ab]

#31 Interferon [ti, ab]

#32 Remdesivir [ti, ab]

#33 GS-5734 [ti, ab]

#34 Oseltamivir [ti, ab]

#35 Lopinavir [ti, ab]

#36 Ritonavir [ti, ab]

#37 Kaletra [ti, ab]

#38 “LPV/r”[ti, ab]

#39 #27-#38/ OR

#40 #26 AND #39

#41 [medline]/lim in #40

#42 #40 NOT #41

### Web of science

#1 TOPIC: “COVID-19”

#2 TOPIC: “SARS-COV-2”

#3 TOPIC: “Novel coronavirus”

#4 TOPIC: “2019-novel coronavirus”

#5 TOPIC: “coronavirus disease-19” [Title/Abstract]

#6 TOPIC: “coronavirus disease 2019” [Title/Abstract]

#7 TOPIC: “COVID19” [Title/Abstract]

#8 TOPIC: “Novel CoV” #9 TOPIC: “2019-nCoV”

#10 TOPIC: “2019-CoV”

#11 TOPIC: “Wuhan-Cov”

#12 TOPIC: “Wuhan Coronavirus”

#13 TOPIC: “Wuhan seafood market pneumonia virus”

#14 TOPIC: “Middle East Respiratory Syndrome”

#15 TOPIC: “MERS”

#16 TOPIC: “MERS-CoV”

#17 TOPIC: “Severe Acute Respiratory Syndrome”

#18 TOPIC: “SARS”

#19 TOPIC: “SARS-CoV”

#20 TOPIC: “SARS-Related”

#21 TOPIC: “SARS-Associated”

#22 #1-#21/OR

#23 TOPIC: (“Antiviral*”)

#24 TOPIC: (“Ribavirin”)

#25 TOPIC: (“Virazole”)

#26 TOPIC: (“Interferon”)

#27 TOPIC: (“Remdesivir”)

#28 TOPIC: (“GS-5734”)

#29 TOPIC: (“Oseltamivir”)

#30 TOPIC: (“Lopinavir”)

#31 TOPIC: (“Ritonavir”)

#32 TOPIC: (“Kaletra”)

#33 TOPIC: (“LPV/r”)

#34 #23-#33/ OR

#35 #22 AND #34

### Cochrane Library

#1 MeSH descriptor: [Middle East Respiratory Syndrome Coronavirus] explode all trees

#2 MeSH descriptor: [Severe Acute Respiratory Syndrome] explode all trees

#3 MeSH descriptor: [SARS Virus] explode all trees

#4 “COVID-19”:ti,ab,kw

#5 “SARS-COV-2”:ti,ab,kw

#6 “Novel coronavirus”:ti,ab,kw

#7 “2019-novel coronavirus” :ti,ab,kw

#8 “Novel CoV” :ti,ab,kw

#9 “2019-nCoV” :ti,ab,kw

#10 “2019-CoV” :ti,ab,kw

#11 “coronavirus disease-19” :ti,ab,kw

#12 “coronavirus disease 2019” :ti,ab,kw

#13 “COVID19” :ti,ab,kw

#14 “Wuhan-Cov” :ti,ab,kw

#15 “Wuhan Coronavirus” :ti,ab,kw

#16 “Wuhan seafood market pneumonia virus” :ti,ab,kw

#17 “Middle East Respiratory Syndrome” :ti,ab,kw

#18 “MERS”:ti,ab,kw

#19 “MERS-CoV”:ti,ab,kw

#20 “Severe Acute Respiratory Syndrome”:ti,ab,kw

#21 “SARS” :ti,ab,kw

#22 “SARS-CoV” :ti,ab,kw

#23 “SARS-Related”:ti,ab,kw

#24 “SARS-Associated”:ti,ab,kw

#25 #1-#24/ OR

#26 MeSH descriptor: [Antiviral agents] explode all trees

#27 (“Antiviral*”): ti, ab, kw

#28 (“Ribavirin”): ti, ab, kw

#29 (“Virazole”): ti, ab, kw

#30 (“Interferon”): ti, ab, kw

#31 (“Remdesivir”): ti, ab, kw

#32 (“GS-5734”): ti, ab, kw

#33 (“Oseltamivir”): ti, ab, kw

#34 (“Lopinavir”): ti, ab, kw

#35 (“Ritonavir”): ti, ab, kw

#36 (“Kaletra”): ti, ab, kw

#37 (“LPV/r”): ti, ab, kw

#38 #26-#37/ OR

#39 #25 AND #38

### CNKI

#1 主题:(“新型冠状病毒”)

#2 主题:(“COVID-19”)

#3 主题:(“SARS-COV-2”)

#4 主题:(“2019-nCoV “)

#5 主题:(“2019-CoV “)

#6 主题:(“武汉冠状病毒”)

#7 主题:(“中东呼吸综合征”)

#8 主题:(“严重急性呼吸综合征”)

#9 主题:(“SARS”)

#10 主题:(“MERS”)

#11 主题:(“MERS-CoV “)

#12 #1-#11/ OR

#13 主题:(“抗病毒”)

#14 主题:(“干扰素”)

#15 主题:(“利巴韦林”)

#16 主题:(“病毒唑”)

#17 主题:(“三氮唑核苷”)

#18 主题:(“尼斯可”)

#19 主题:(“瑞德西韦”)

#20 主题:(“奥司他韦”)

#21 主题:(“达菲”)

#22 主题:(“特敏福”)

#23 主题:(“克流感”)

#24 主题:(“洛匹那韦”)

#25 主题:(“利托那?”)

#26 主题:(“利托纳韦”)

#27 主题:(“克力芝”)

#28 #13-#27/ OR

#29 #12 AND #28

### CBM

#1 “新型冠状病毒”[常用字段:智能]

#2 “COVID-19”[常用字段:智能]

#3 “SARS-COV-2”[常用字段:智能]

#4 “2019-nCoV”[常用字段:智能]

#5 “2019-CoV”[常用字段:智能]

#6 “武汉冠状病毒”[常用字段:智能]

#7 “中东呼吸综合征冠状病毒”[不加权:扩展]

#8 “中东呼吸综合征”[常用字段:智能]

#9 “MERS”[常用字段:智能]

#10 “MERS-CoV”[常用字段:智能]

#11 “严重急性呼吸综合征”[不加权:扩展]

#12 “SARS 病毒”[不加权:扩展]

#13 “严重急性呼吸综合征”

#14 “SARS”[常用字段:智能]

#15 #1-#14 / OR

#16 “抗病毒药” [不加权:扩展]

#17 “抗病毒”[常用字段:智能]

#18 “干扰素”[常用字段:智能]

#19 “利巴韦林”[常用字段:智能]

#20 “病毒唑”[常用字段:智能]

#21 “三氮唑核苷”[常用字段:智能]

#22 “尼斯可”[常用字段:智能]

#23 “瑞德西韦”[常用字段:智能]

#24 “奥司他韦”[常用字段:智能]

#25 “达菲”[常用字段:智能]

#26 “特敏福”[常用字段:智能]

#27 “克流感”[常用字段:智能]

#28 “洛匹那韦”[常用字段:智能]

#29 “利托那韦”[常用字段:智能]

#30 “克力芝”[常用字段:智能]

#31 #16-#30/ OR

#32 #15 AND #31

### Wanfang

#1 主题:(“新型冠状病毒”)

#2 主题:(“COVID-19”)

#3 主题:(“SARS-COV-2”)

#4 主题:(“2019-nCoV “)

#5 主题:(“2019-CoV “)

#6 主题:(“武汉冠状病毒”)

#7 主题:(“中东呼吸综合征”)

#8 主题:(“严重急性呼吸综合征”)

#9 主题:(“SARS”)

#10 主题:(“MERS”)

#11 主题:(“MERS-CoV “)

#12 #1-#11/ OR

#13 主题:(“抗病毒”)

#14 主题:(“干扰素”)

#15 主题:(“利巴韦林”)

#16 主题:(“病毒唑”)

#17 主题:(“三氮唑核苷”)

#18 主题:(“尼斯可”)

#19 主题:(“瑞德西韦”)

#20 主题:(“奥司他韦”)

#21 主题:(“达菲”)

#22 主题:(“特敏福”)

#23 主题:(“克流感”)

#24 主题:(“洛匹那韦”)

#25 主题:(“利托那韦”)

#26 主题:(“利托纳韦”)

#27 主题:(“克力芝 “)

#28 #13-#27/ OR

#29 #12 AND #28

## Supplementary Material 2. Data extraction (Table 1-8)

### Lopinavir/ ritonavir (LPV/r)

**Table 1.**
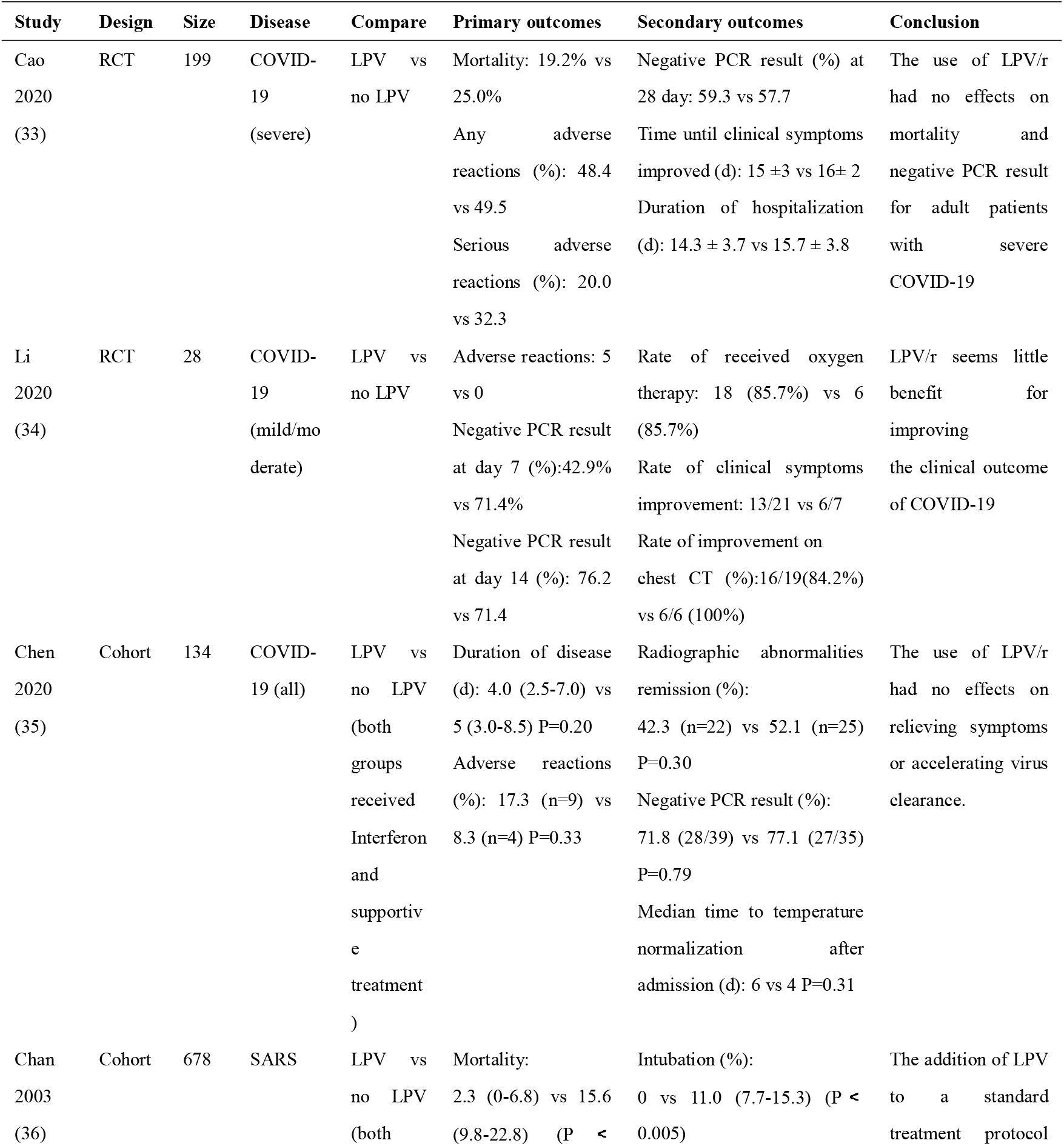
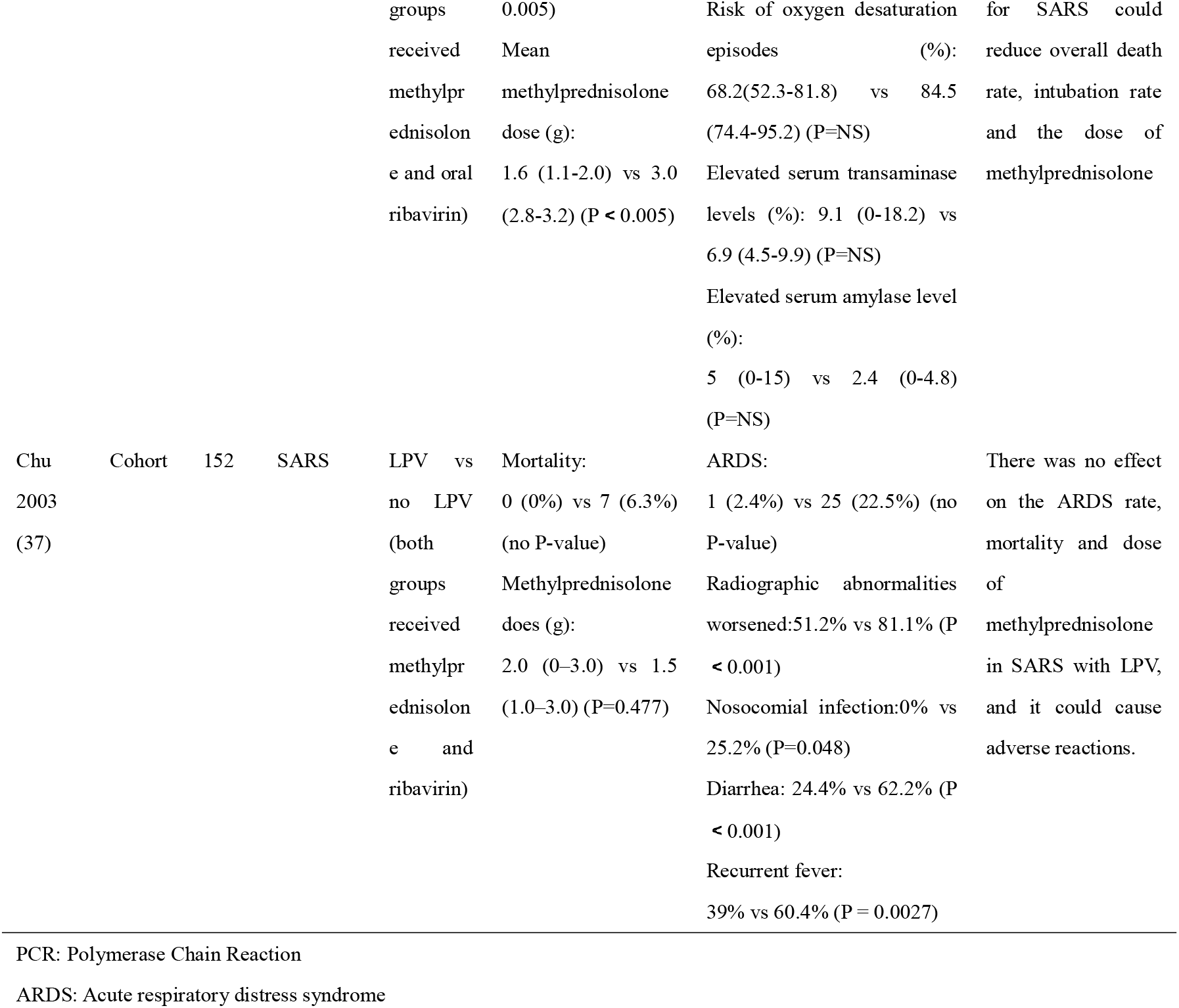
Evidence summary of LPV/r

### Arbidol

**Table 2.** Evidence summary of Arbidol

| Study | Design | Size | Disease | Compare | Primary outcomes | Secondary outcomes | Conclusion |
| --- | --- | --- | --- | --- | --- | --- | --- |
| Chen<br>2020<br>(35) | Cohort | 82 | COVID-19 | Arbidol vs<br>no Arbidol<br>(both<br>groups<br>received<br>Interferon<br>and<br>supportive<br>treatment) | Duration of disease<br>(d): 3.5 (2.0-6.0) vs<br>5 (3.0-8.5) P=0.20<br>Adverse reactions<br>(%)<br>8.8 (n=3) vs 8.3<br>(n=4) P=0.33 | Radiographic<br>abnormalities remission<br>(%):<br>35.3 (n=12) vs 52.1<br>(n=25) P=0.30<br>Negative PCR result (%)<br>82.6 (19/23) vs 77.1<br>(27/35) P=0.79<br>Median time to<br>temperature<br>normalization after<br>admission (d):<br>6 vs 4 P=0.31 | The use of Arbidol had<br>no effects on relieving<br>symptoms or<br>accelerating virus<br>clearance. |
| Deng<br>2020<br>(38) | Cohort | 33 | COVID-19 | Arbidol plus LPV/r vs no LPV/r | Negative PCR result (%):15 /16 (94%) vs 9/17 (52.9%) | Improvement of chest CT scans: 11/16 (69%) vs 5/17 (29%) | Arbidol plus LPV/r could increase the rate of negative PCR and chest improvement |
| Li<br>2020<br>(34) | RCT | 23 | COVID-19 | Arbidol vs no Arbidol | Adverse reactions: 0 vs 0<br>Negative PCR result at day 7 (%):62.5 vs 71.4<br>Negative PCR result at day 14 (%): 87.5 vs 71.4 | Rate of received oxygen therapy: 11 (68.8%) vs 6 (85.7%)<br>Rate of clinical symptoms improvement: 14/16 vs 6/7<br>Rate of improvement on chest CT (%):10/15(66.7%) vs 6/6 (100%) | Arbidol seems little benefit for improving the clinical outcome of COVID-19 |

PCR: Polymerase Chain Reaction

### Interferon (IFN)

**Table 3.**
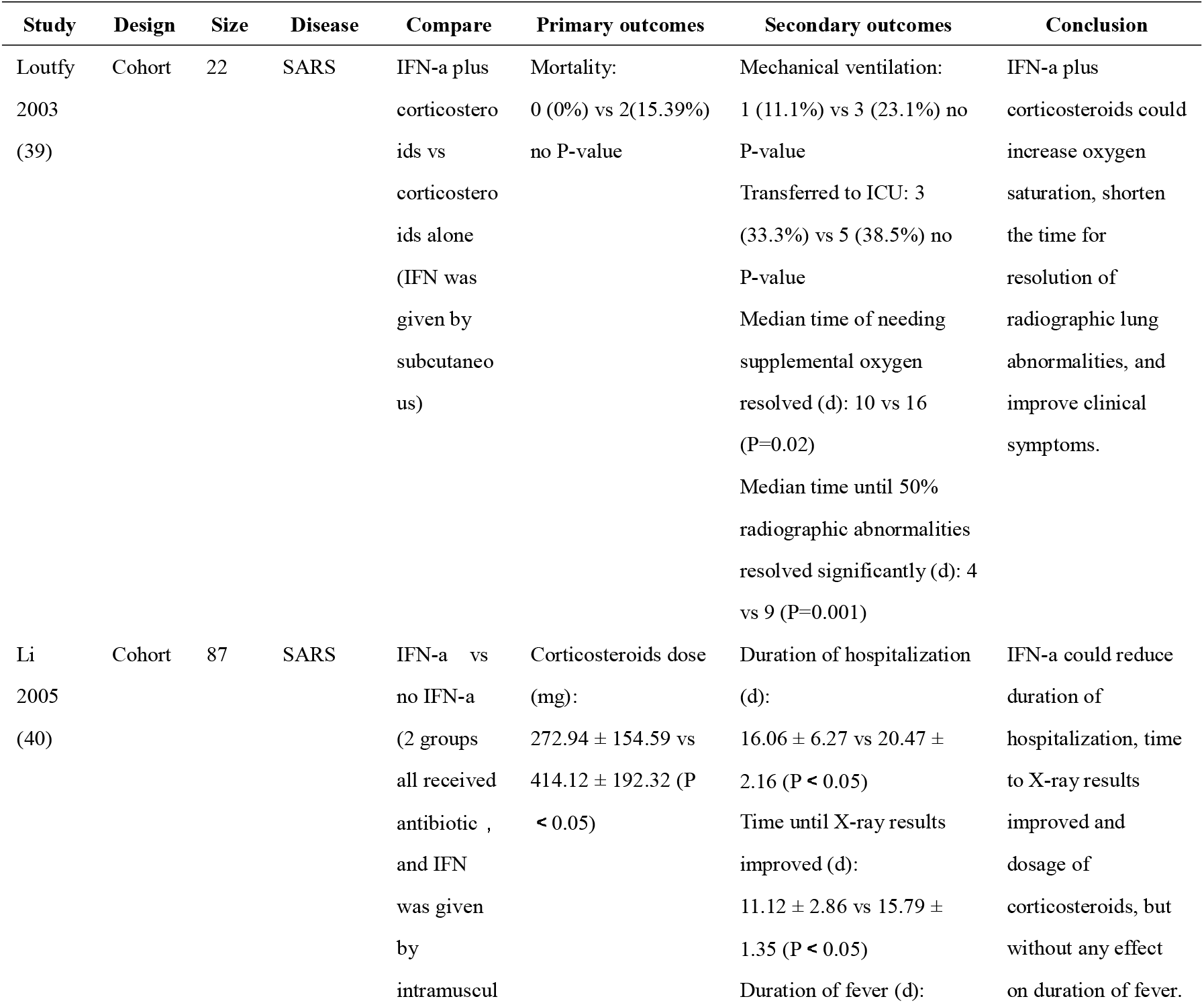
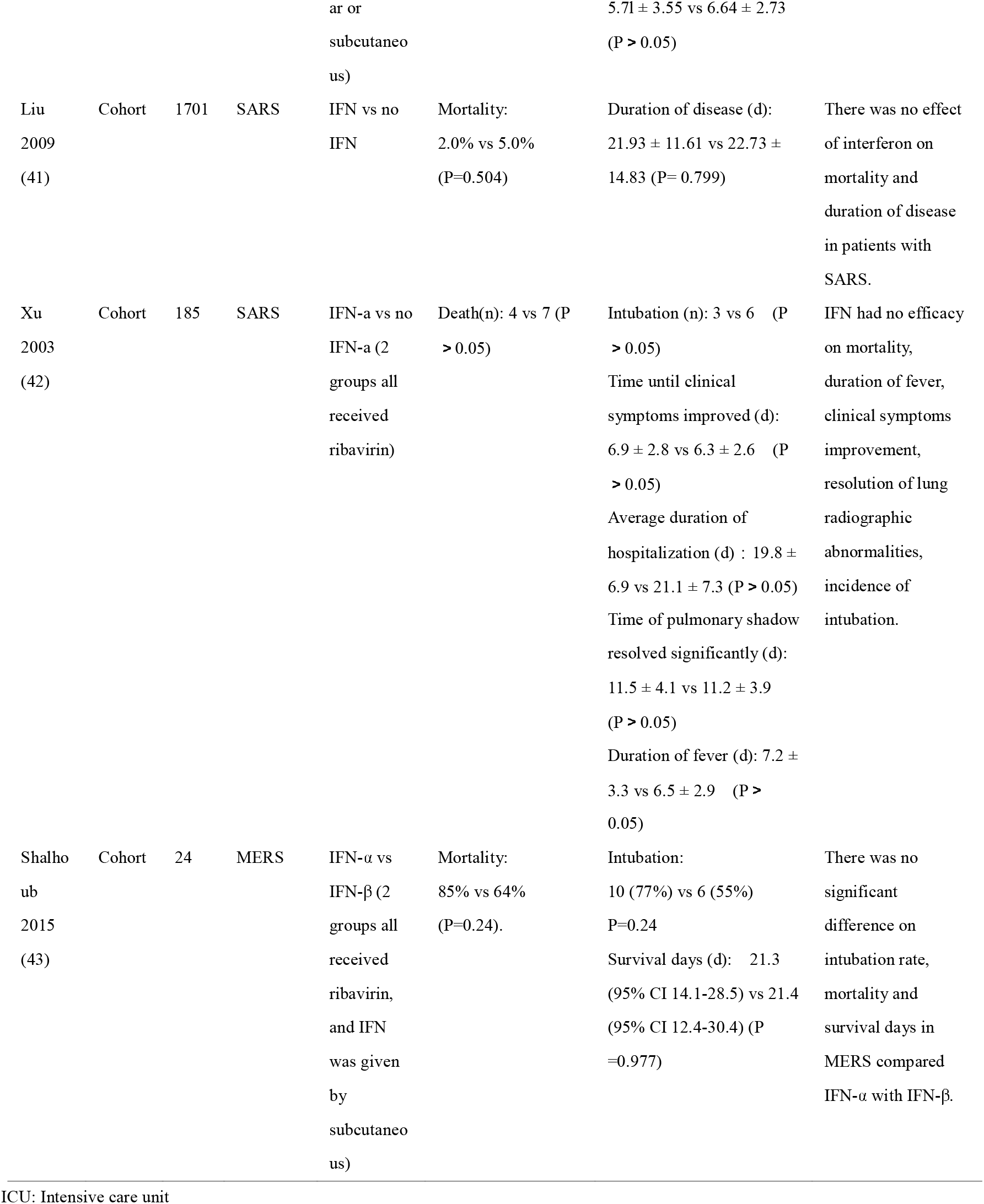
Evidence summary of IFN

### Ribavirin (RBV)

**Table 4.** Evidence summary of RBV

| Study | Design | Size | Disease | Compare | Primary outcomes | Secondary outcomes | Conclusion |
| --- | --- | --- | --- | --- | --- | --- | --- |
| Leong<br>2004 (44) | Cohort | 229 | SARS | RBV vs<br>no RBV | Crude death rate:<br>10 (10.3%) vs 17<br>(12.9%) (P =0.679) | Admitted to ICU:<br>19 (19.6%) vs 27<br>(20.5%) (P >0.999) | There was no effect of<br>ribavirin on crude<br>death rate, numbers of |
|  |  |  |  |  | Anemia:<br>24 (24.7%) vs 27<br>(20.5%) (P=0.521) | Myocardial injury:<br>3 (3.1%) vs 4 (3.0%)<br>(P > 0.999) | admitted to ICU, and<br>adverse reaction in<br>patients with SARS. |
| Chiou<br>2005 (45) | Cohort | 51 | SARS | RBV vs<br>no RBV | Hypoxemia:<br>39% vs 14% (P<br>=0.398)<br>Anemia:<br>73% vs 14% (P=<br>0.006) | Peak CRP level (mg/dL):<br>10.3 ± 11.6 vs 5.8 ± 6.2<br>(P > 0.05)<br>Peak LDH level (IU/L):<br>392.8 ± 307.5 vs 162.5 ±<br>98.0 (P=0.017) | Ribavirin could<br>increase LDH levels<br>and risk of anemia in<br>SARS, but had no<br>effect on CRP level<br>and the occurrence of<br>hypoxemia. |
| Lau 2009<br>(46) | Cohort | 1104 | SARS | RBV vs<br>no RBV | Mortality (%):<br>Hong Kong: 17.0<br>(95% CI 6.5-27.8)<br>vs 15.4 (95% CI<br>13.2-17.6) (P=0.77);<br>Toronto: 13.4 (95%<br>CI 0-33.0) vs 16.6<br>(95% CI 0-44.9)<br>(P=0.85) | None | Early treatment of<br>SARS with ribavirin<br>had no effect on<br>mortality. |
| Wang<br>2005 (47) | Cohort | 90 | SARS | RBV vs<br>no RBV | Duration of<br>corticosteroids use<br>(d):<br>21.5 ± 7.4 vs 2 7.1 ±<br>3.8 (P< 0.0 1) | None | Ribavirin could reduce<br>the duration of<br>corticosteroids use, but<br>the time length of<br>ribavirin was not<br>associated with the<br>reduction. |
| Muller<br>2007 (48) | Cohort | 306 | SARS | RBV vs<br>no RBV | Death:<br>20 (11%) vs 10<br>(8%) (P=0.42)<br>Hemolytic anemia:<br>57% vs 30% (P<br><0.0001) | Mechanical ventilation:<br>15% vs 15% (P=0.88)<br>Bradycardia:<br>34% vs 17% (P=0.0009)<br>Hyperamylasemia:<br>11% vs 3% (P=0.032)<br>Hypocalcemia:<br>55% vs 38% (P=0.0038)<br>Hypomagnesemia:<br>50% vs 5% (P<0.0001) | High-dose ribavirin<br>was associated with<br>serious adverse<br>reactions in SARS, but<br>had no effect on<br>mortality. |
| Liu 2009<br>(41) | Cohort | 1702 | SARS | RBV vs<br>no RBV | Mortality:<br>4.4% vs 5.4%<br>(P=0.340) | Duration of disease<br>(d):<br>23.23±14.45 vs 22.19 ±<br>15.01 (P= 0.044) | Ribavirin cannot<br>reduce the mortality,<br>but prolong the<br>duration of disease. |

CRP: C-reactive protein
LDH: Lactate dehydrogenase
ICU: Intensive care unit

### Oseltamivir

**Table 5.**
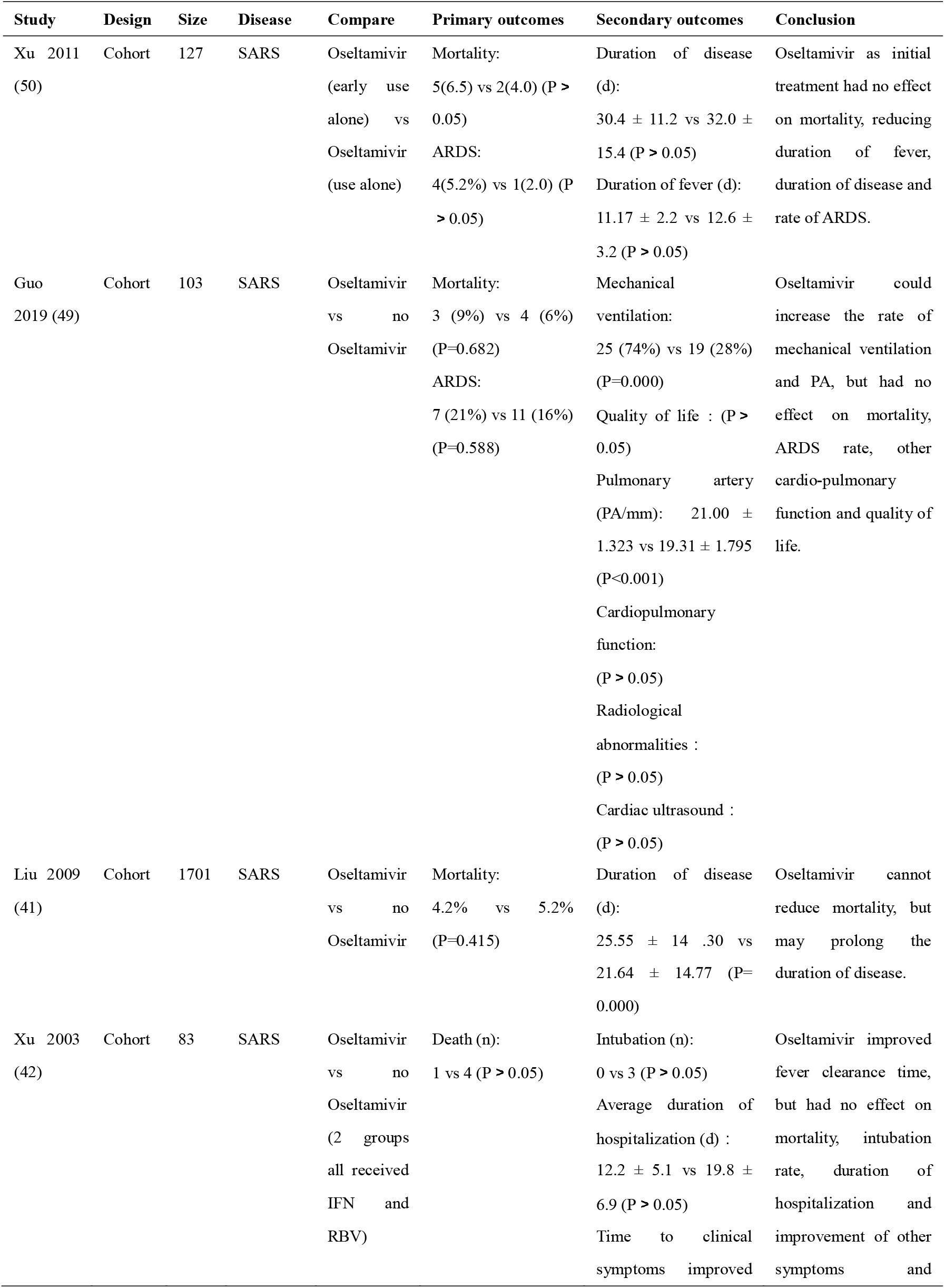
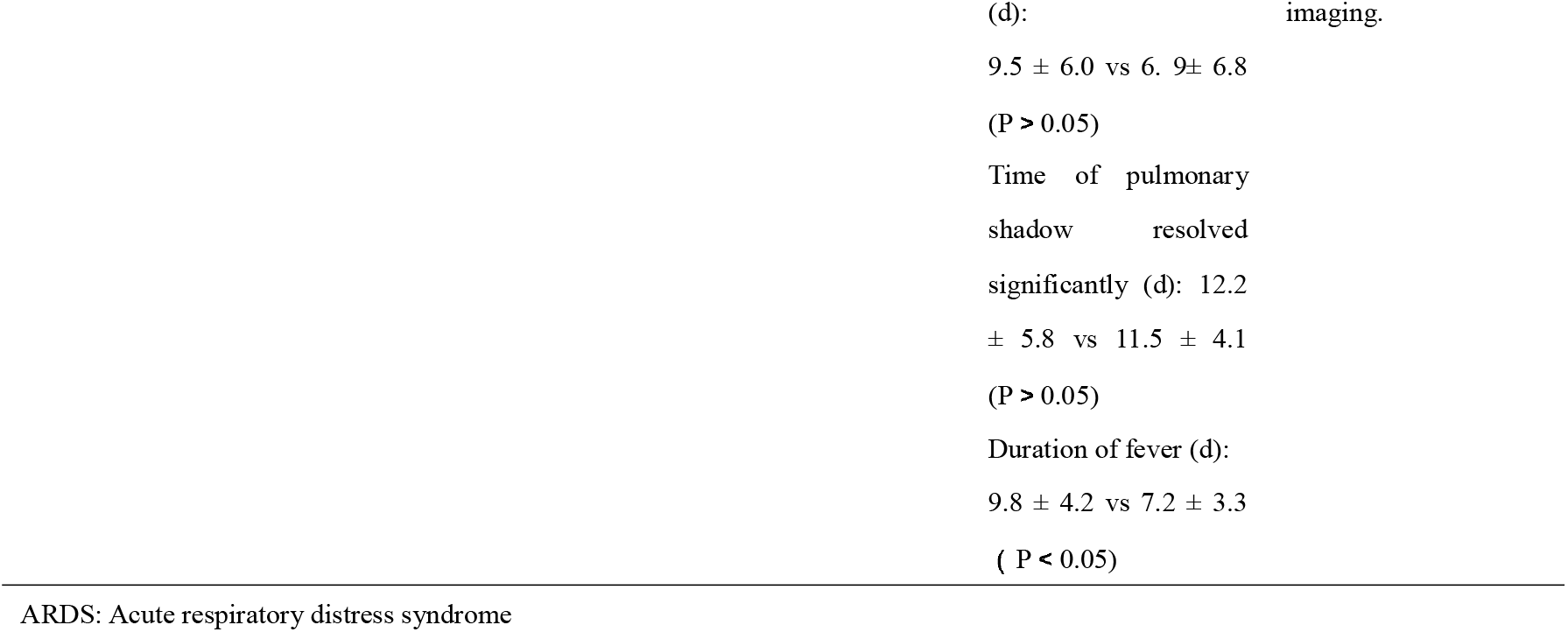
Evidence summary of Oseltamivir

### Combination of IFN and RBV

**Table 6.**
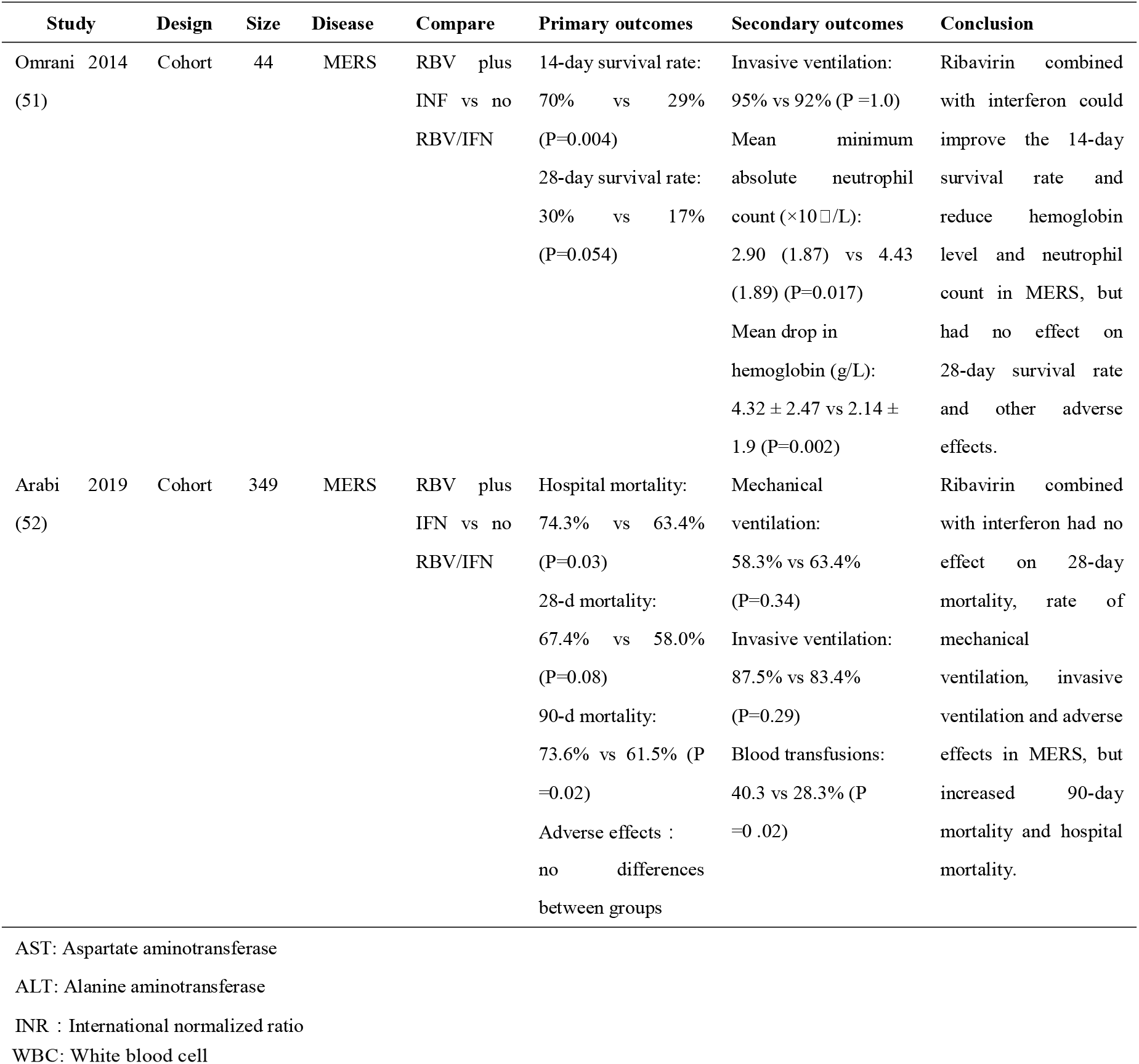
Evidence summary of Combination

### Favipiravir

**Table 7.** Evidence summary of Favipiravir

| Study | Design | Size | Disease | Compare | Primary outcomes | Secondary outcomes | Conclusion |
| --- | --- | --- | --- | --- | --- | --- | --- |
| Chen 2020 (53)<sup>b</sup> | RCT | 236 | COVID-19 | Favipiravir vs Arbidol | clinical recovery rate of day 7: 61.21% (71/116) vs 51.67% (62/120)<br>Adverse reactions: 37/116 vs 28/120 | auxiliary oxygen therapy or noninvasive mechanical ventilation rate: 22.5% (27/120) vs 18.1% (21/116)<br>new dyspnea: 4/116 vs 14/120<br>respiratory failure: 4/116 vs 4/120 | favipiravir can be considered as a preferred treatment for moderate COVID-19 |

### Hydroxychloroquine (HCQ)

**Table 8.** Evidence summary of HCQ

| Study | Design | Size | Disease | Compare | Primary outcomes | Secondary outcomes | Conclusion |
| --- | --- | --- | --- | --- | --- | --- | --- |
| Chen 2020 (54) | RCT | 30 | COVID-19 | Hydroxychloroquine vs none (two groups all received IGN) | Rate of virological cured at 7 days: 86.7 vs 93.3<br>Adverse reactions: 4 (26.7%) vs 3 (20.0%) $P > 0.05$ | Time until negative result (d): 4 (1-9) vs 2 (1-4) $P > 0.05$<br>Duration of fever (d): 1(0-2) vs 1(0-3) | Hydroxychloroquine has little benefit for COVID-19 patients |
| Chen 2020 (55) | RCT | 62 | COVID-19 | Hydroxychloroquine vs none | Adverse reactions: 2 vs 0 | Duration of fever (d): 2.2 ± 0.4 vs 3.2 ± 1.3<br>Rate of improved pneumonia: 80.6%, (25/31) vs 54.8% (17 of 31)<br>Incidence of clinical symptom improvement: 31 vs 27 | HCQ can be considered as a preferred treatment for moderate COVID-19 |

## Supplementary Material 3. GRADE evidence profile (Table 1-8)

**Table 1.**
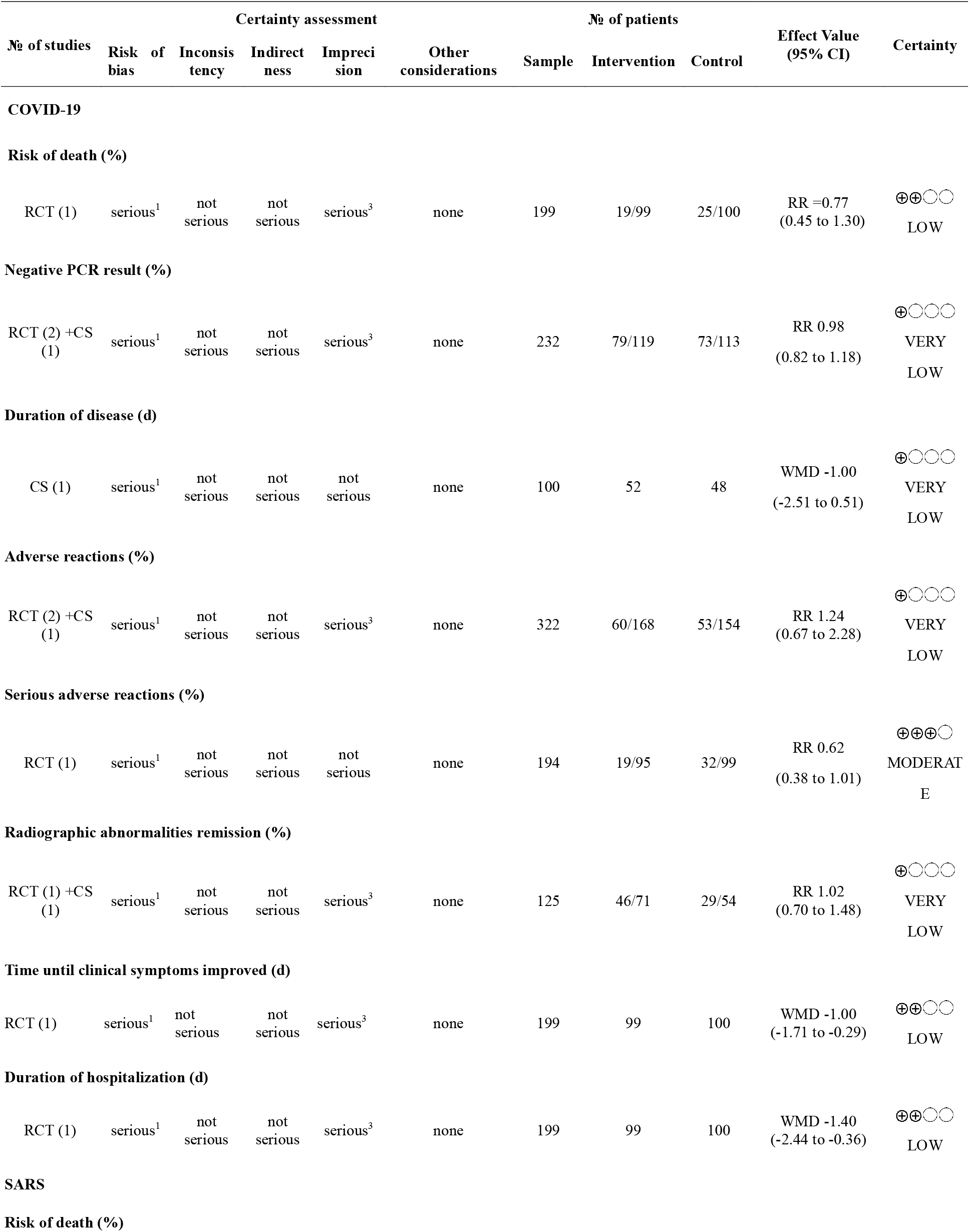
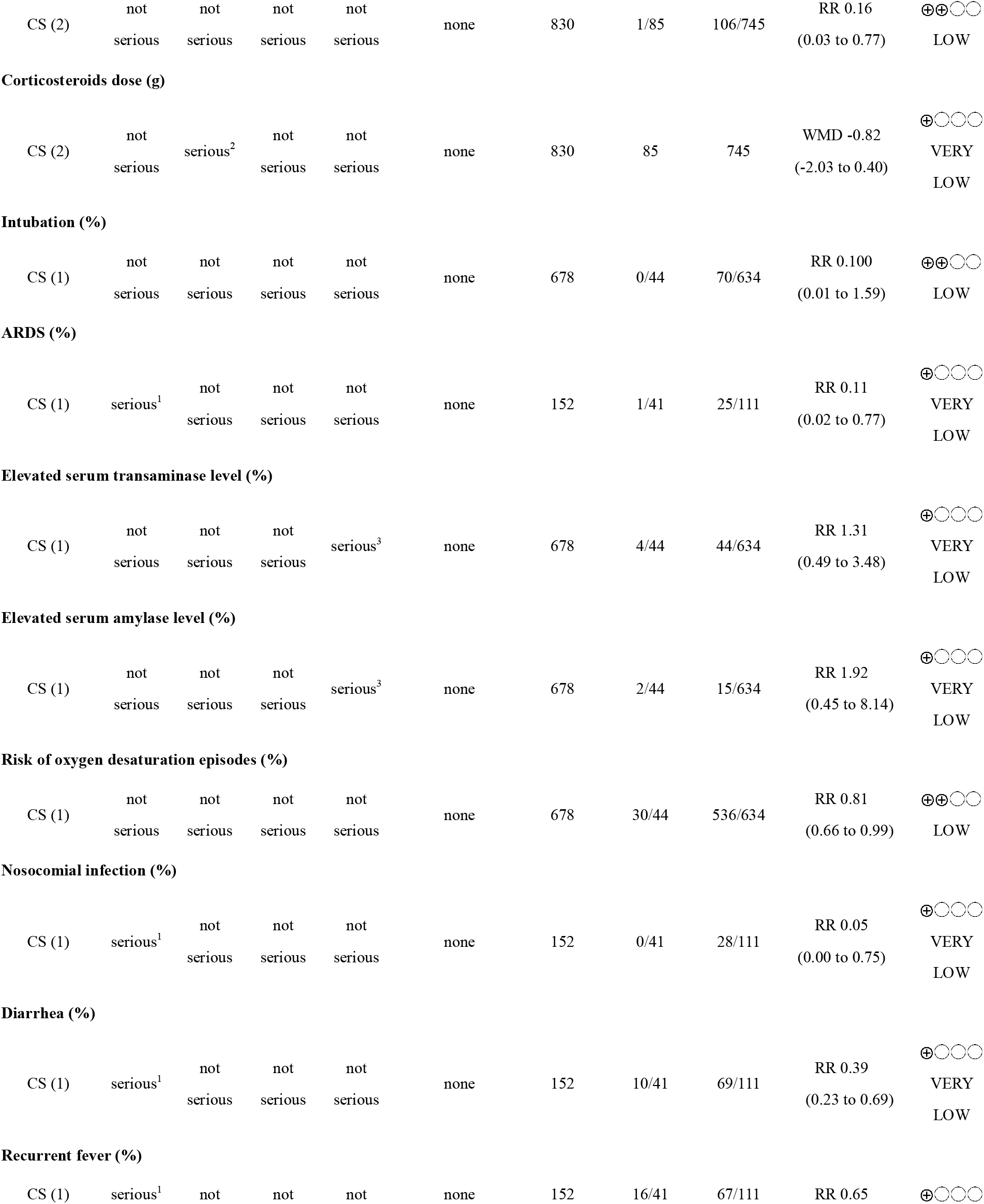
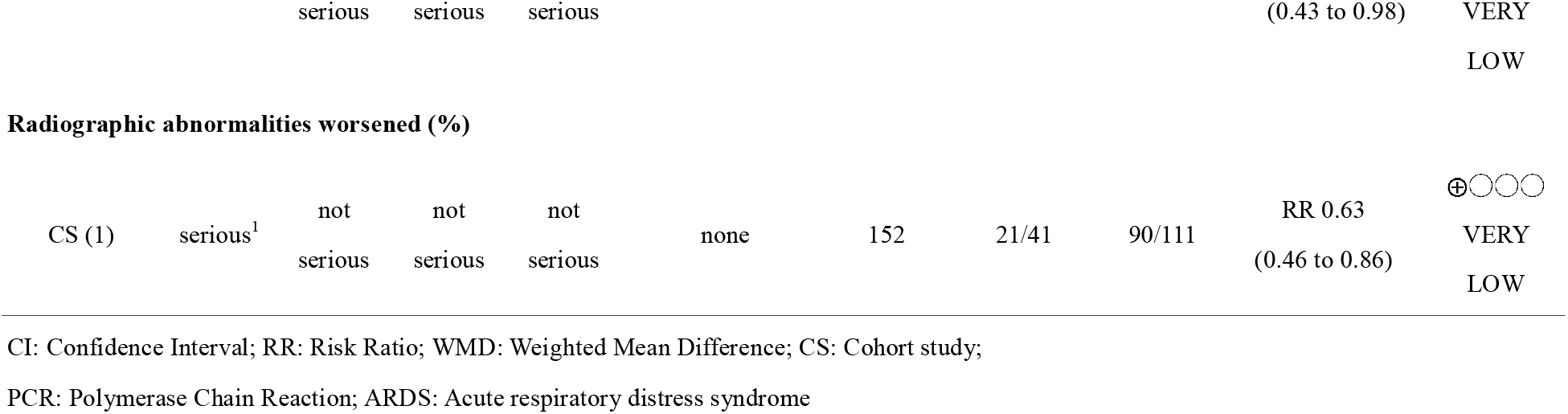
lopinavir/ ritonavir (LPV/r)

**Table 2.** Arbidol

| № of studies | Certainty assessment |  |  |  |  | № of patients |  |  | Effect Value (95% CI) | Certainty |
| --- | --- | --- | --- | --- | --- | --- | --- | --- | --- | --- |
|  | Risk of bias | Inconsistency | Indirectness | Imprecision | Other considerations | Sample | Intervention | Control |  |  |
| COVID-19 |  |  |  |  |  |  |  |  |  |  |
| Duration of disease (d) |  |  |  |  |  |  |  |  |  |  |
| CS (1) | not serious | not serious | not serious | serious<sup>3</sup> | none | 82 | 34 | 48 | WMD -1.70 (-3.28 to -0.12) | ⊕○○○<br>VERY LOW |
| Negative PCR result (%) |  |  |  |  |  |  |  |  |  |  |
| RCT (1) + CS (2) | not serious | serious<sup>2</sup> | not serious | not serious | none | 114 | 48/55 | 41/59 | RR 1.27 (0.93 to 1.73) | ⊕○○○<br>VERY LOW |
| Adverse reactions (%) |  |  |  |  |  |  |  |  |  |  |
| CS (1) | not serious | not serious | not serious | serious<sup>3</sup> | none | 82 | 3/34 | 4/48 | RR 1.06 (0.25 to 4.43) | ⊕○○○<br>VERY LOW |
| Radiographic abnormalities remission (%) |  |  |  |  |  |  |  |  |  |  |
| RCT (1) + CS (2) | not serious | not serious | not serious | serious<sup>3</sup> | none | 136 | 43/65 | 34/71 | RR 1.23 (0.63 to 2.40) | ⊕○○○<br>VERY LOW |
| Incidence of receiving oxygen therapy (%) |  |  |  |  |  |  |  |  |  |  |
| RCT (1) | serious<sup>1</sup> | not serious | not serious | serious<sup>3</sup> | none | 23 | 11/16 | 6/7 | RR 0.80 (0.51 to 1.26) | ⊕⊕⊕○<br>MODERATE |
| Incidence of clinical symptoms improvement |  |  |  |  |  |  |  |  |  |  |
| RCT (1) | serious<sup>1</sup> | not serious | not serious | serious<sup>3</sup> | none | 23 | 14/16 | 6/7 | RR 1.02 (0.72 to 1.46) | ⊕⊕⊕○<br>MODERATE |

**Table 3.**
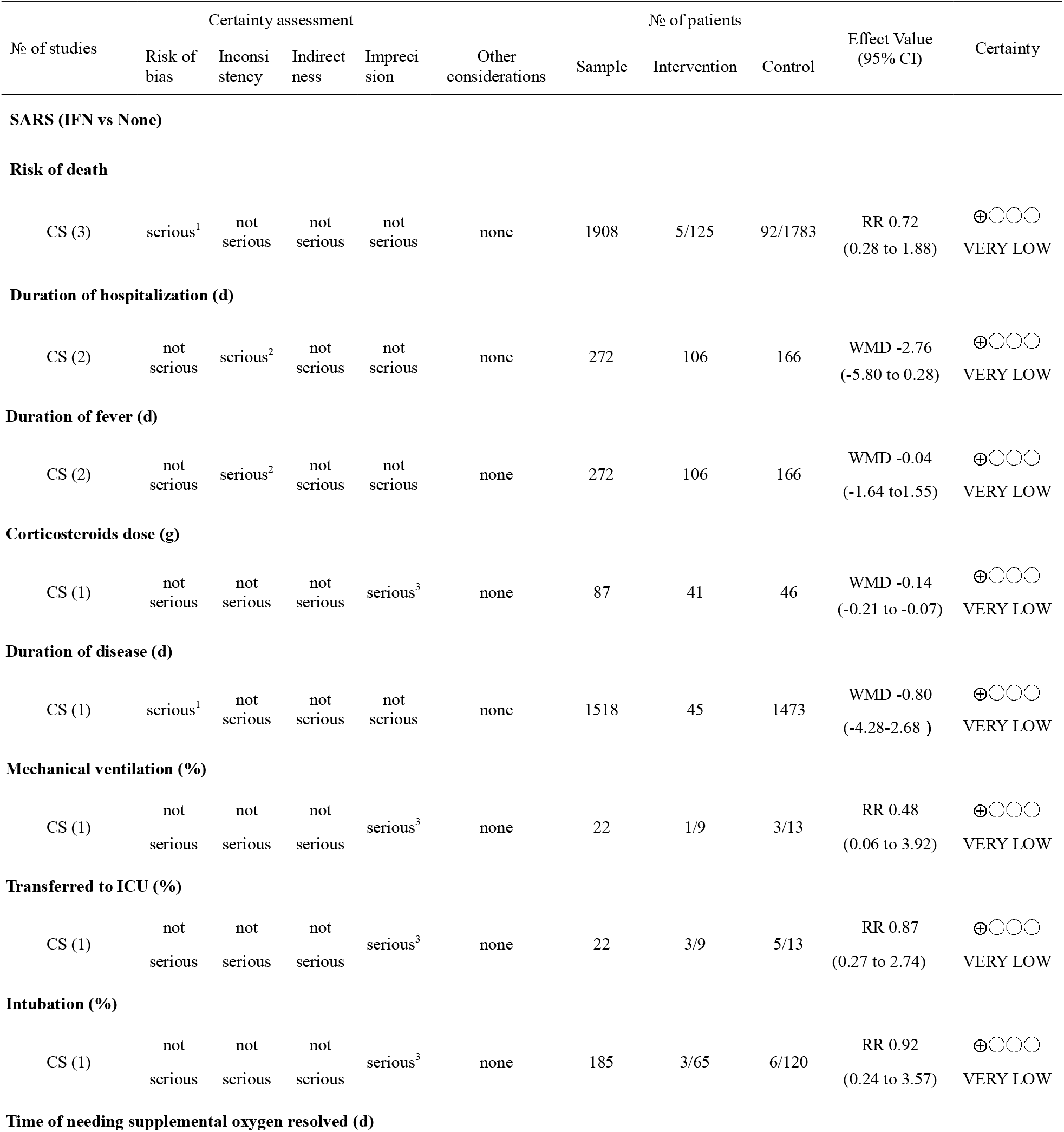
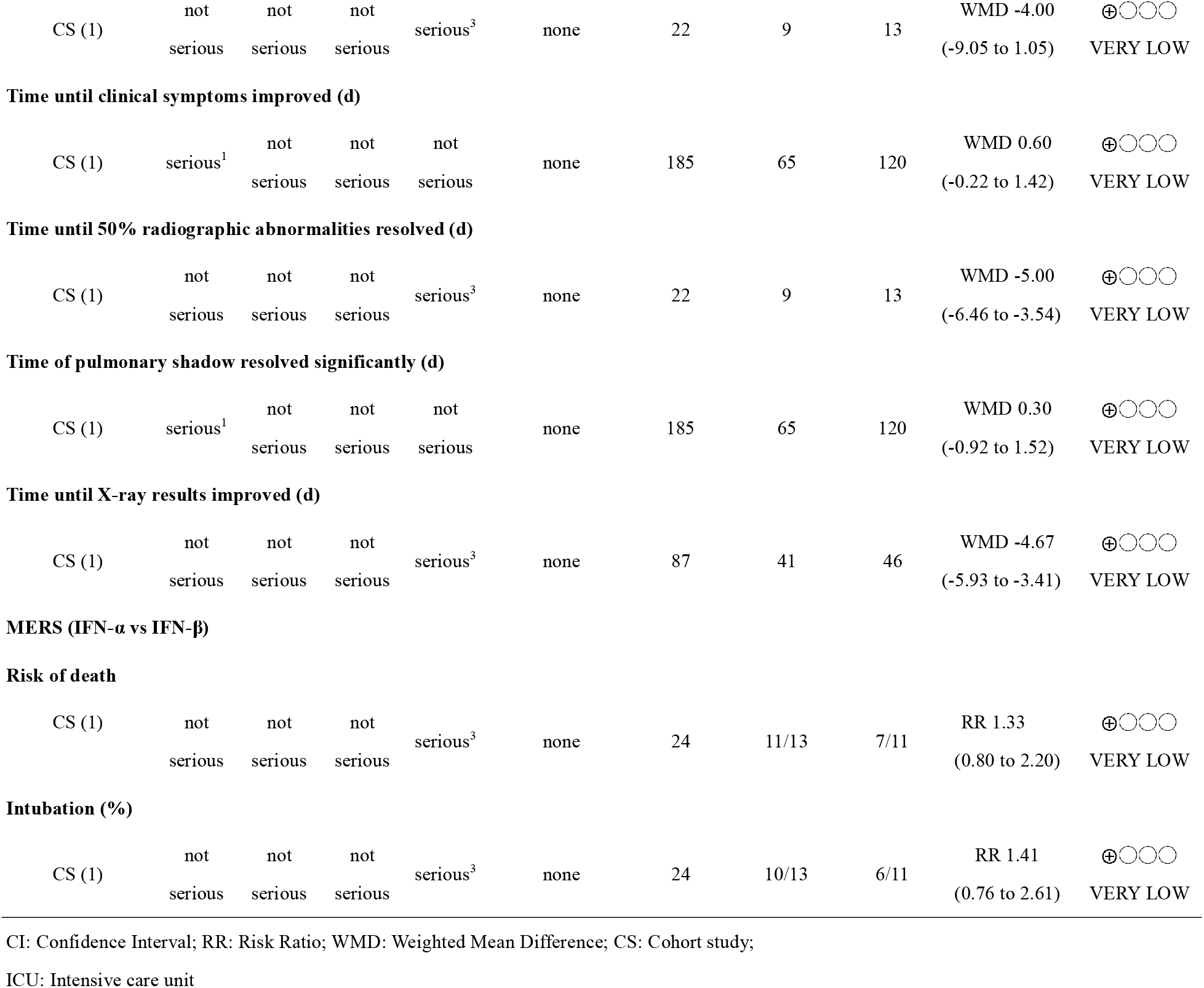
Interferon (IFN)

**Table 4.**
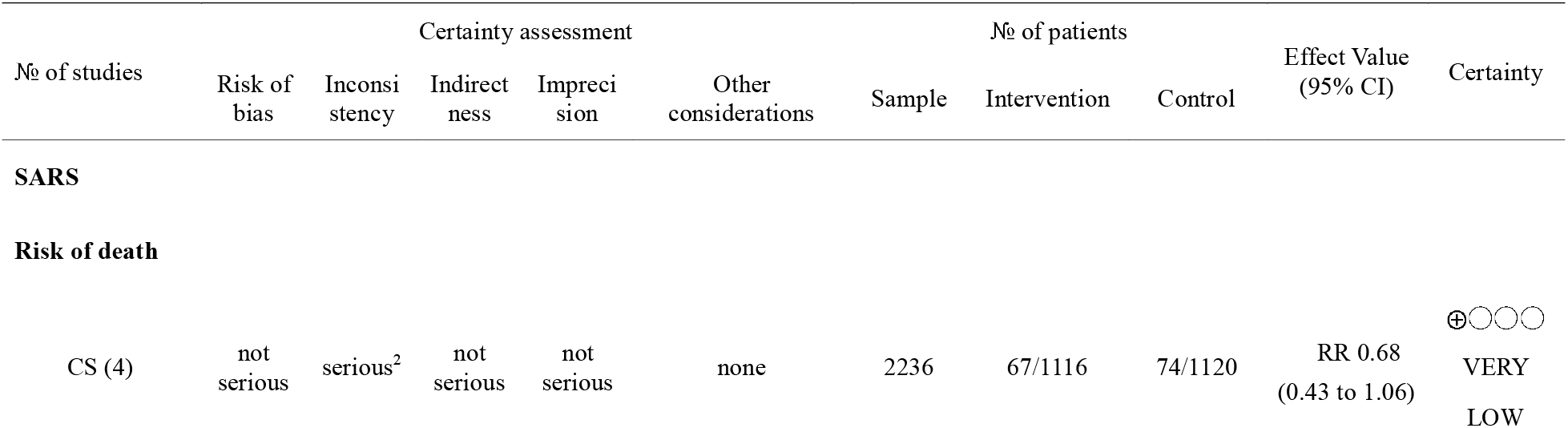
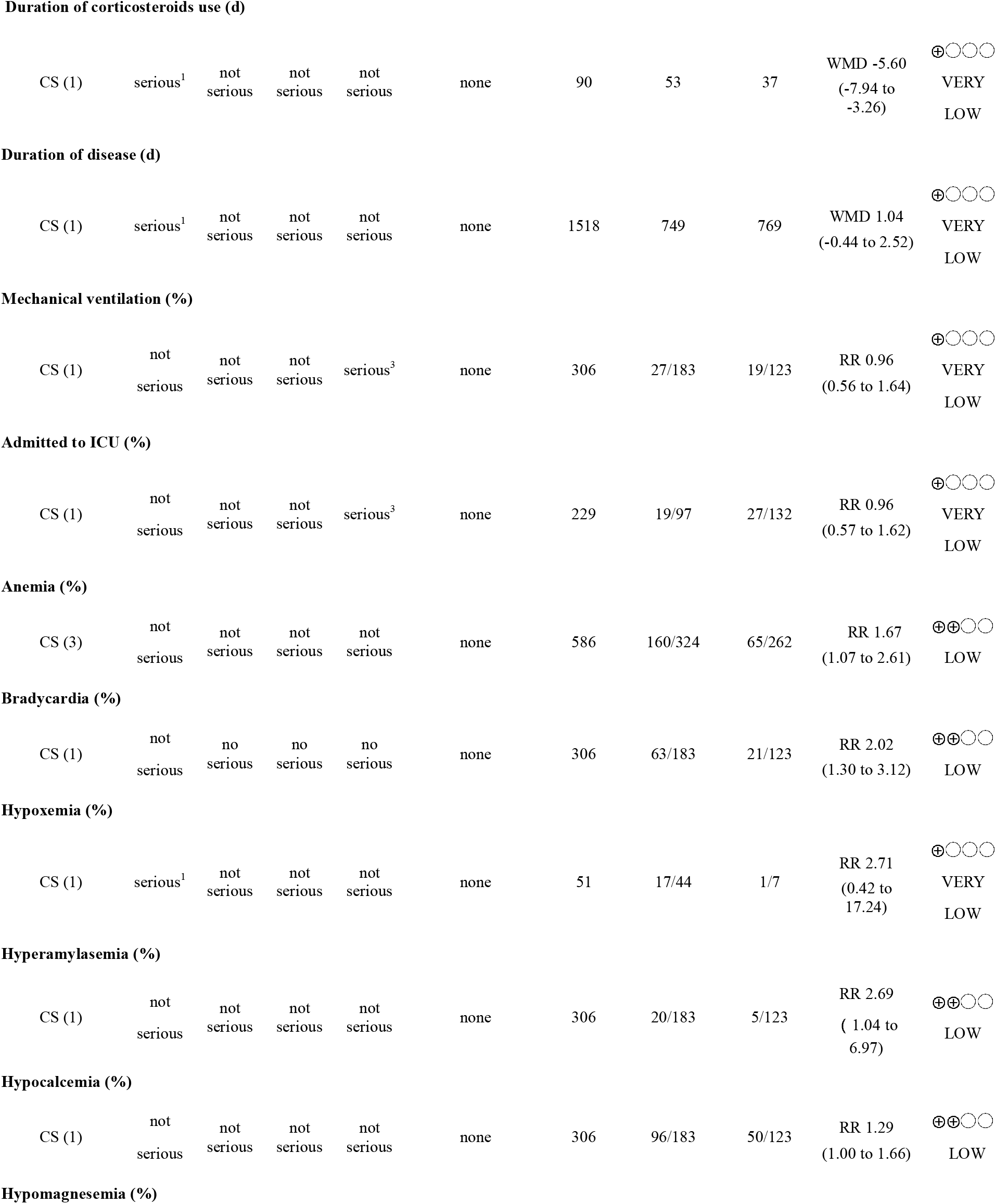
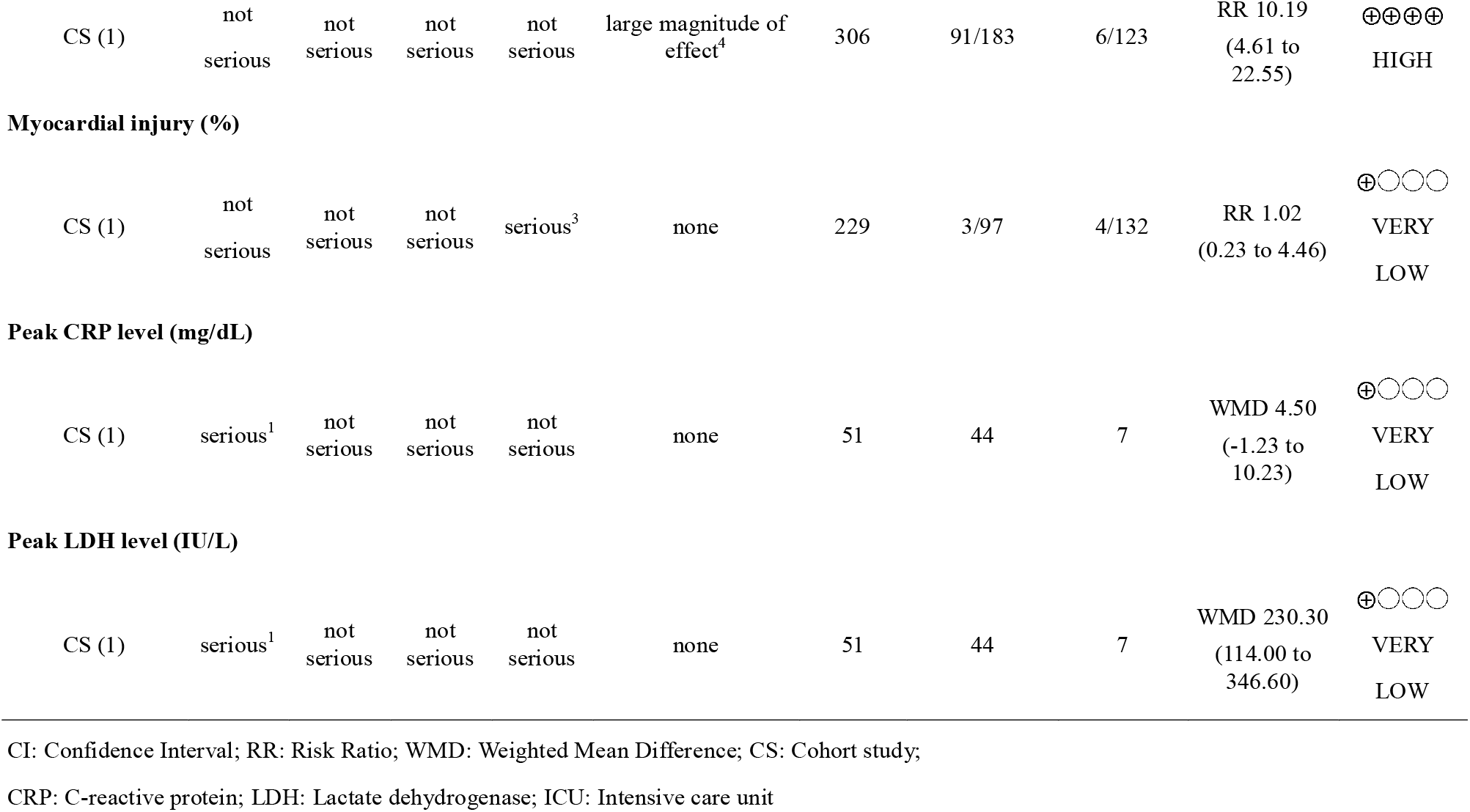
Ribavirin (RBV)

**Table 5.**
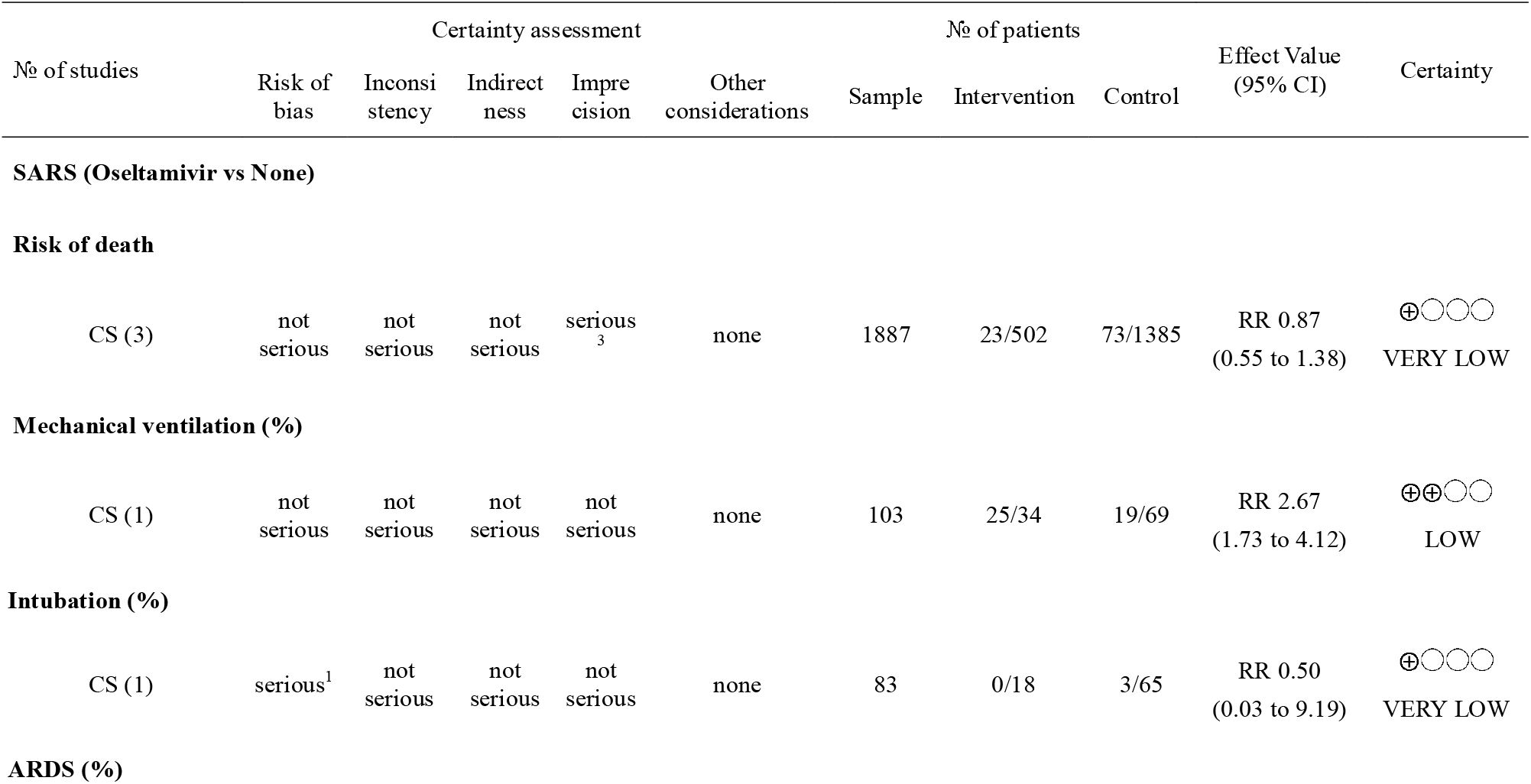
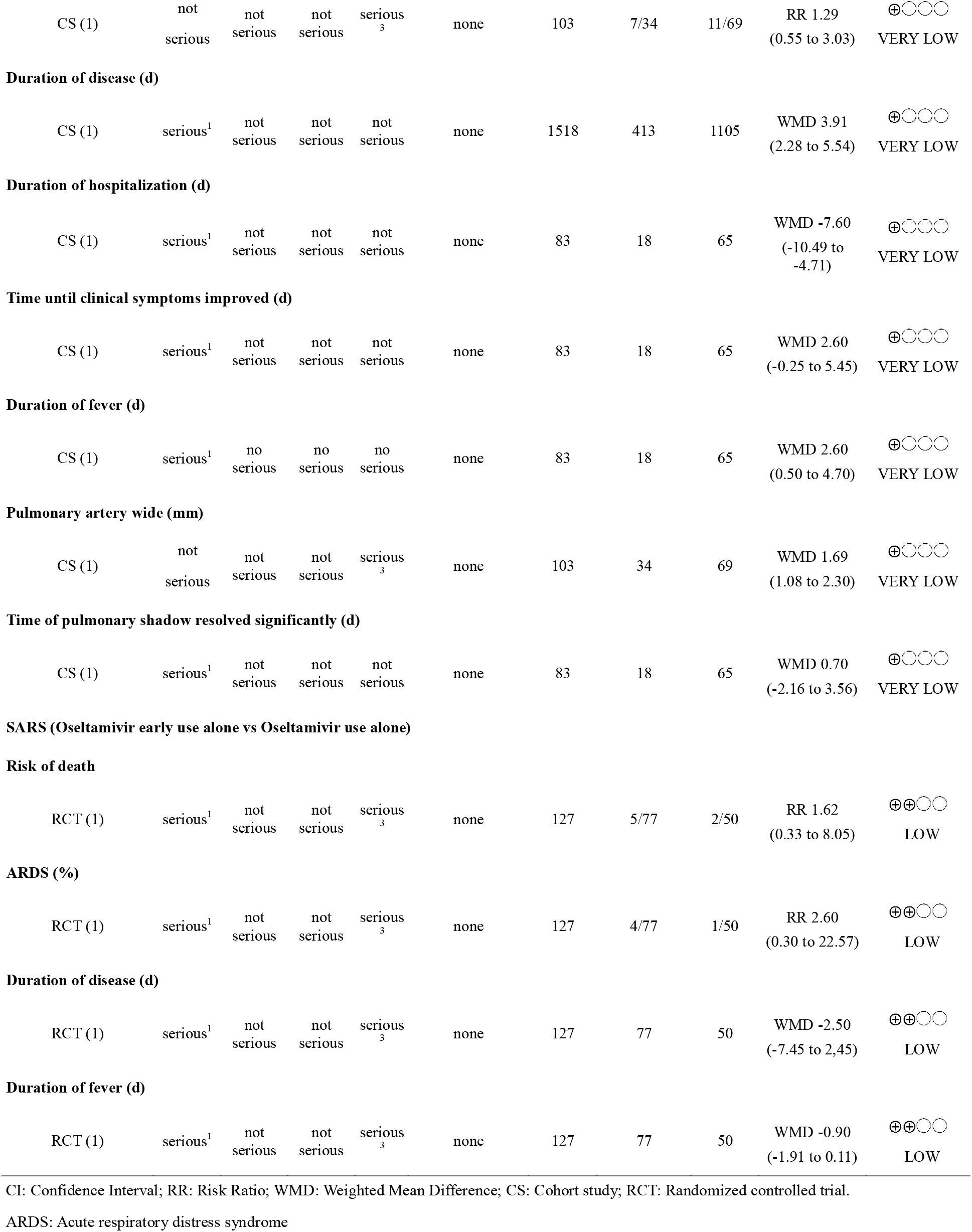
Oseltamivir

| № of studies | Certainty assessment |  |  |  |  | № of patients |  |  | Effect Value<br>(95% CI) | Certainty |
| --- | --- | --- | --- | --- | --- | --- | --- | --- | --- | --- |
|  | Risk of bias | Inconsistency | Indirectness | Imprecision | Other considerations | Sample | Intervention | Control |  |  |
| SARS (Oseltamivir vs None) |  |  |  |  |  |  |  |  |  |  |
| Risk of death |  |  |  |  |  |  |  |  |  |  |
| CS (3) | not serious | not serious | not serious | serious<sub>3</sub> | none | 1887 | 23/502 | 73/1385 | RR 0.87<br>(0.55 to 1.38) | ⊕○○○<br>VERY LOW |
| Mechanical ventilation (%) |  |  |  |  |  |  |  |  |  |  |
| CS (1) | not serious | not serious | not serious | not serious | none | 103 | 25/34 | 19/69 | RR 2.67<br>(1.73 to 4.12) | ⊕⊕○○<br>LOW |
| Intubation (%) |  |  |  |  |  |  |  |  |  |  |
| CS (1) | serious<sup>1</sup> | not serious | not serious | not serious | none | 83 | 0/18 | 3/65 | RR 0.50<br>(0.03 to 9.19) | ⊕○○○<br>VERY LOW |
| ARDS (%) |  |  |  |  |  |  |  |  |  |  |
|  | Risk of<br>bias | Inconsistency | Indirectness | Imprecision | Other<br>considerations | Sample | Intervention | Control |  |  |
| CS (1) | not<br>serious | not<br>serious | not<br>serious | serious<sub>3</sub> | none | 103 | 7/34 | 11/69 | RR 1.29<br>(0.55 to 3.03) | ⊕○○○<br>VERY LOW |
| **Duration of disease (d)** |  |  |  |  |  |  |  |  |  |  |
| CS (1) | serious<sup>1</sup> | not<br>serious | not<br>serious | not<br>serious | none | 1518 | 413 | 1105 | WMD 3.91<br>(2.28 to 5.54) | ⊕○○○<br>VERY LOW |
| **Duration of hospitalization (d)** |  |  |  |  |  |  |  |  |  |  |
| CS (1) | serious<sup>1</sup> | not<br>serious | not<br>serious | not<br>serious | none | 83 | 18 | 65 | WMD -7.60<br>(-10.49 to -4.71) | ⊕○○○<br>VERY LOW |
| **Time until clinical symptoms improved (d)** |  |  |  |  |  |  |  |  |  |  |
| CS (1) | serious<sup>1</sup> | not<br>serious | not<br>serious | not<br>serious | none | 83 | 18 | 65 | WMD 2.60<br>(-0.25 to 5.45) | ⊕○○○<br>VERY LOW |
| **Duration of fever (d)** |  |  |  |  |  |  |  |  |  |  |
| CS (1) | serious<sup>1</sup> | no<br>serious | no<br>serious | no<br>serious | none | 83 | 18 | 65 | WMD 2.60<br>(0.50 to 4.70) | ⊕○○○<br>VERY LOW |
| **Pulmonary artery wide (mm)** |  |  |  |  |  |  |  |  |  |  |
| CS (1) | not<br>serious | not<br>serious | not<br>serious | serious<sub>3</sub> | none | 103 | 34 | 69 | WMD 1.69<br>(1.08 to 2.30) | ⊕○○○<br>VERY LOW |
| **Time of pulmonary shadow resolved significantly (d)** |  |  |  |  |  |  |  |  |  |  |
| CS (1) | serious<sup>1</sup> | not<br>serious | not<br>serious | not<br>serious | none | 83 | 18 | 65 | WMD 0.70<br>(-2.16 to 3.56) | ⊕○○○<br>VERY LOW |
| **SARS (Oseltamivir early use alone vs Oseltamivir use alone)** |  |  |  |  |  |  |  |  |  |  |
| **Risk of death** |  |  |  |  |  |  |  |  |  |  |
| RCT (1) | serious<sup>1</sup> | not<br>serious | not<br>serious | serious<sub>3</sub> | none | 127 | 5/77 | 2/50 | RR 1.62<br>(0.33 to 8.05) | ⊕⊕○○<br>LOW |
| **ARDS (%)** |  |  |  |  |  |  |  |  |  |  |
| RCT (1) | serious<sup>1</sup> | not<br>serious | not<br>serious | serious<sub>3</sub> | none | 127 | 4/77 | 1/50 | RR 2.60<br>(0.30 to 22.57) | ⊕⊕○○<br>LOW |
| **Duration of disease (d)** |  |  |  |  |  |  |  |  |  |  |
| RCT (1) | serious<sup>1</sup> | not<br>serious | not<br>serious | serious<sub>3</sub> | none | 127 | 77 | 50 | WMD -2.50<br>(-7.45 to 2.45) | ⊕⊕○○<br>LOW |
| **Duration of fever (d)** |  |  |  |  |  |  |  |  |  |  |
| RCT (1) | serious<sup>1</sup> | not<br>serious | not<br>serious | serious<sub>3</sub> | none | 127 | 77 | 50 | WMD -0.90<br>(-1.91 to 0.11) | ⊕⊕○○<br>LOW |

| № of studies | Certainty assessment |  |  |  |  | № of patients |  |  | Effect Value<br>(95% CI) | Certainty |
| --- | --- | --- | --- | --- | --- | --- | --- | --- | --- | --- |
|  | Risk of<br>bias | Inconsi-<br>stency | Indirect<br>ness | Impreci-<br>sion | Other<br>considerations | Sample | Intervention | Control |  |  |

CI: Confidence Interval; RR: Risk Ratio; WMD: Weighted Mean Difference; CS: Cohort study; RCT: Randomized controlled trial.
ARDS: Acute respiratory distress syndrome

**Table 6.** Ribavirin (RBV) plus Interferon (IFN)

| № of studies | Certainty assessment |  |  |  |  | № of patients |  |  | Effect Value<br>(95% CI) | Certainty |
| --- | --- | --- | --- | --- | --- | --- | --- | --- | --- | --- |
|  | Risk of<br>bias | Inconsi-<br>stency | Indirect<br>ness | Impreci-<br>sion | Other<br>considerations | Sample | Intervention | Control |  |  |
| MERS |  |  |  |  |  |  |  |  |  |  |
| Risk of death |  |  |  |  |  |  |  |  |  |  |
| CS (2) | not<br>serious | serious<sup>2</sup> | not<br>serious | not<br>serious | none | 393 | 120/164 | 146/229 | RR 1.04<br>(0.74 to 1.46) | ⊕○○○<br>VERY<br>LOW |
| Invasive ventilation (%) |  |  |  |  |  |  |  |  |  |  |
| CS (2) | not<br>serious | not<br>serious | not<br>serious | serious<sup>3</sup> | none | 393 | 145/164 | 193/229 | RR 1.05<br>(0.97 to 1.13) | ⊕○○○<br>VERY<br>LOW |
| Mechanical ventilation (%) |  |  |  |  |  |  |  |  |  |  |
| CS (1) | not<br>serious | not<br>serious | not<br>serious | serious<sup>3</sup> | none | 349 | 84/144 | 130/205 | RR 0.92<br>(0.77 to 1.09) | ⊕○○○<br>VERY<br>LOW |
| Blood transfusions (%) |  |  |  |  |  |  |  |  |  |  |
| CS (1) | not<br>serious | not<br>serious | not<br>serious | not<br>serious | none | 349 | 58/144 | 58/205 | RR 1.42<br>(1.06 to 1.91) | ⊕⊕○○<br>LOW |
| Mean drop in haemoglobin (g/L) |  |  |  |  |  |  |  |  |  |  |
| CS (1) | not<br>serious | not<br>serious | not<br>serious | serious<sup>3</sup> | none | 44 | 20 | 24 | WMD 2.18<br>(0.86 to 3.50) | ⊕○○○<br>VERY<br>LOW |
| Mean minimum absolute neutrophil count (×10<sup>9</sup>/L) |  |  |  |  |  |  |  |  |  |  |
| CS (1) | not<br>serious | not<br>serious | not<br>serious | serious<sup>3</sup> | none | 44 | 20 | 24 | WMD -1.43<br>(-2.55 to<br>-0.32) | ⊕○○○<br>VERY<br>LOW |

CI: Confidence Interval; RR: Risk Ratio; WMD: Weighted Mean Difference; CS: Cohort study;

**Table 7.**
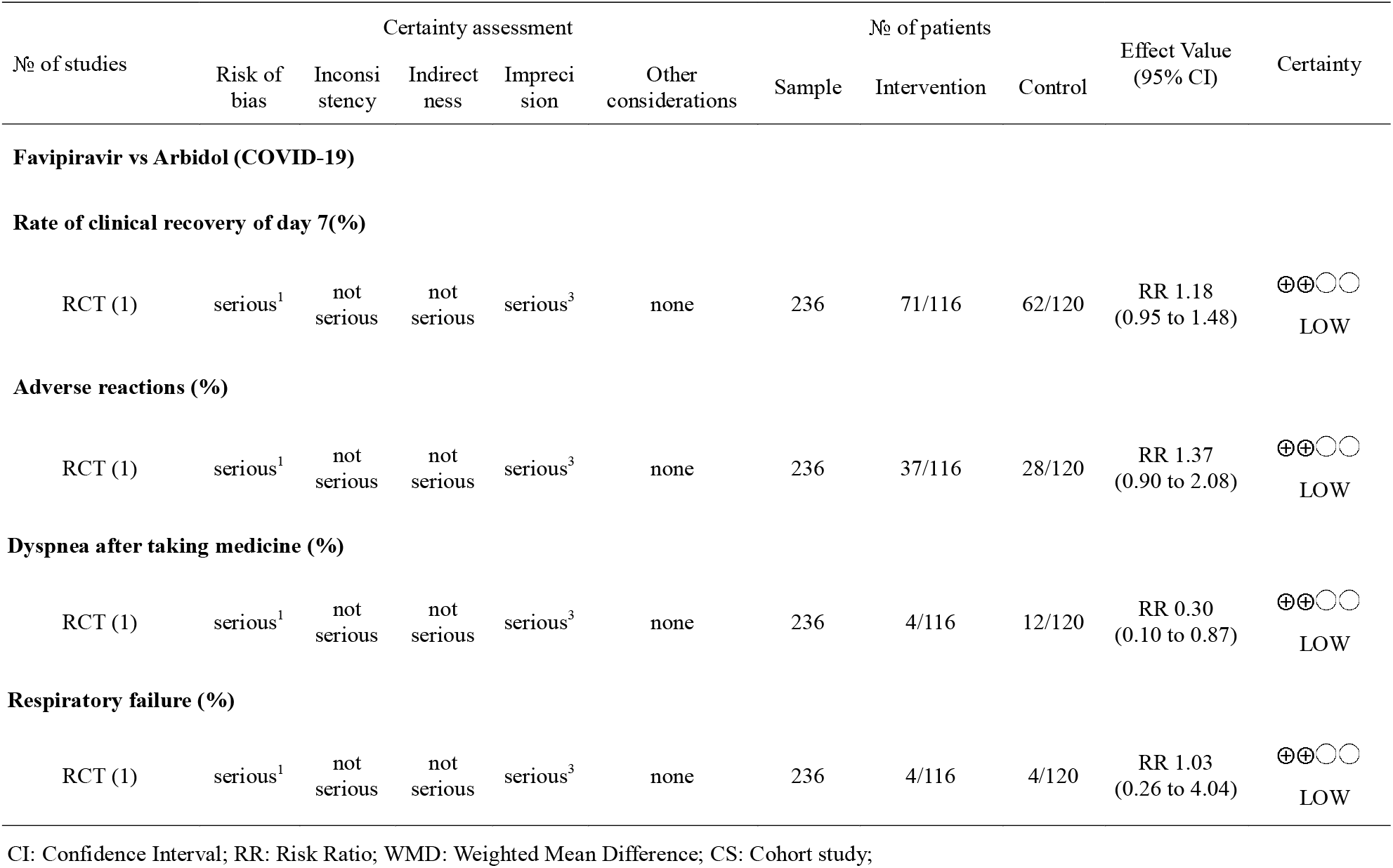
Favipiravir

| № of studies | Certainty assessment |  |  |  |  | № of patients |  |  | Effect Value<br>(95% CI) | Certainty |
| --- | --- | --- | --- | --- | --- | --- | --- | --- | --- | --- |
|  | Risk of<br>bias | Inconsistency | Indirectness | Imprecision | Other considerations | Sample | Intervention | Control |  |  |
| **Respiratory failure (%)** |  |  |  |  |  |  |  |  |  |  |
| RCT (1) | serious<sup>1</sup> | not serious | not serious | serious<sup>3</sup> | none | 236 | 4/116 | 4/120 | RR 1.03<br>(0.26 to 4.04) | ⊕⊕○○<br>LOW |

CI: Confidence Interval; RR: Risk Ratio; WMD: Weighted Mean Difference; CS: Cohort study;

**Table 8.**
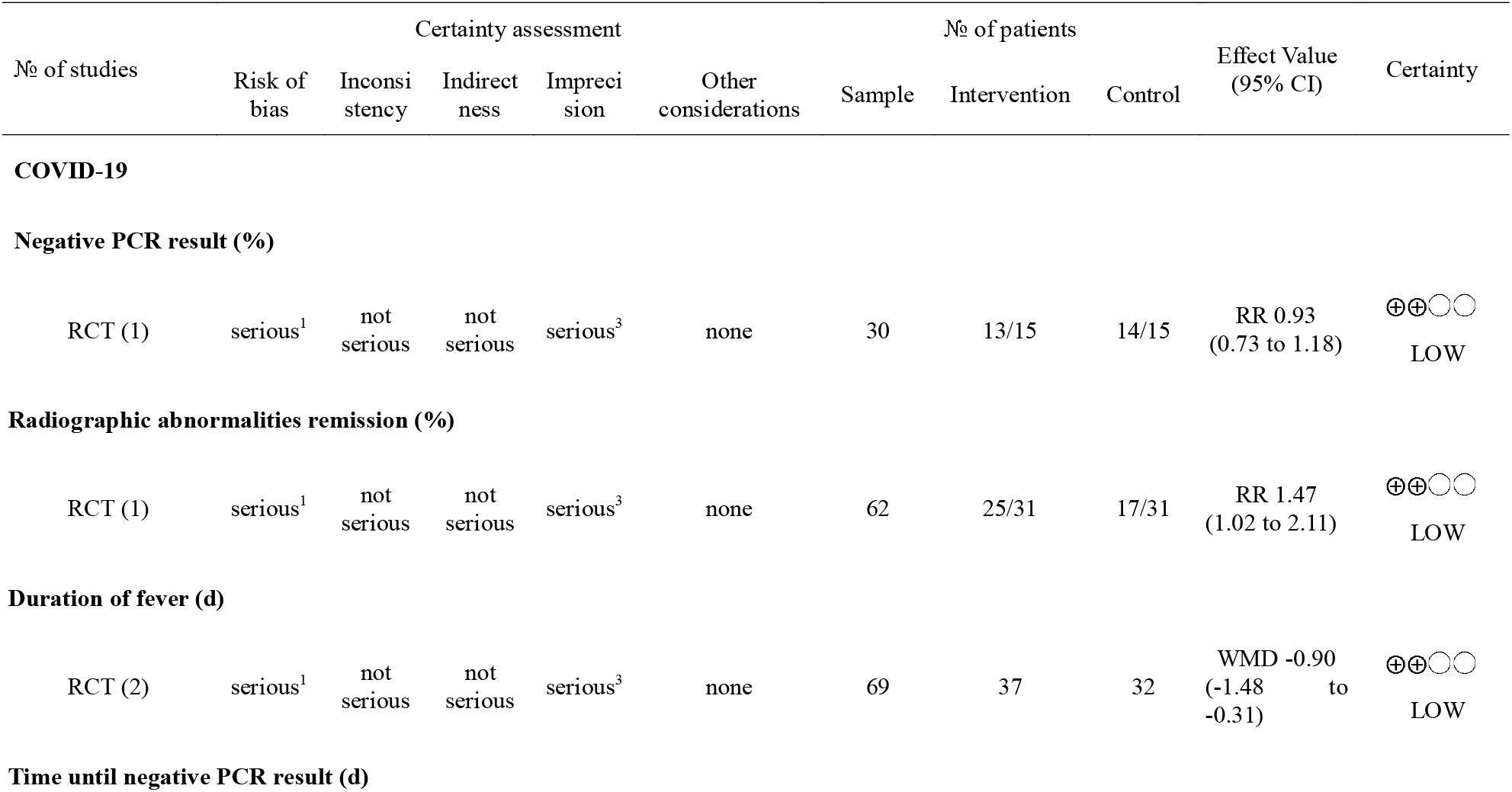
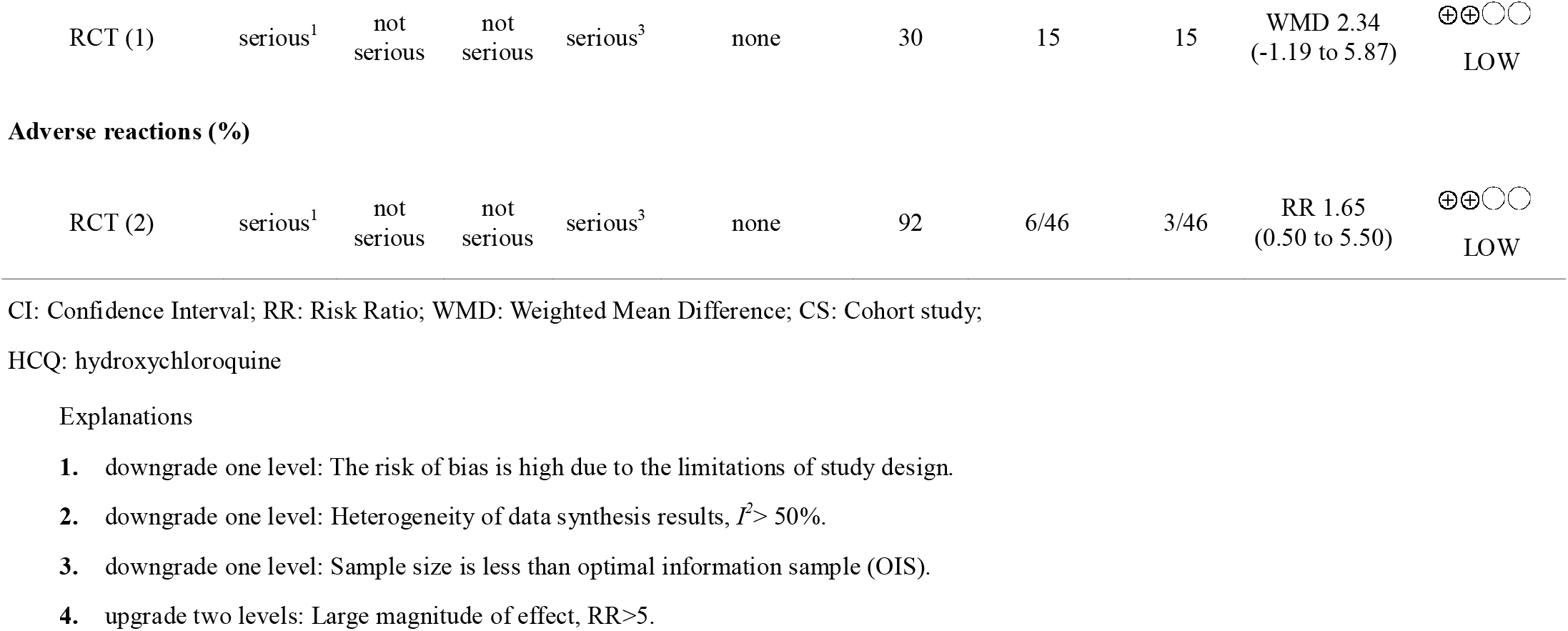
HCQ

| № of studies | Certainty assessment |  |  |  |  | № of patients |  |  | Effect Value<br>(95% CI) | Certainty |
| --- | --- | --- | --- | --- | --- | --- | --- | --- | --- | --- |
|  | Risk of<br>bias | Inconsistency | Indirectness | Imprecision | Other considerations | Sample | Intervention | Control |  |  |
| RCT (2) | serious<sup>1</sup> | not serious | not serious | serious<sup>3</sup> | none | 69 | 37 | 32 | WMD -0.90<br>(-1.48 to -0.31) | ⊕⊕○○<br>LOW |
|  | Risk of bias | Inconsistency | Indirectness | Imprecision | Other considerations | Sample | Intervention | Control |  |  |
| RCT (1) | serious<sup>1</sup> | not serious | not serious | serious<sup>3</sup> | none | 30 | 15 | 15 | WMD 2.34<br>(-1.19 to 5.87) | ⊕⊕○○<br>LOW |
| **Adverse reactions (%)** |  |  |  |  |  |  |  |  |  |  |
| RCT (2) | serious<sup>1</sup> | not serious | not serious | serious<sup>3</sup> | none | 92 | 6/46 | 3/46 | RR 1.65<br>(0.50 to 5.50) | ⊕⊕○○<br>LOW |

CI: Confidence Interval; RR: Risk Ratio; WMD: Weighted Mean Difference; CS: Cohort study;
HCQ: hydroxychloroquine

